# Fronto-limbic neural variability as a transdiagnostic correlate of emotion dysregulation

**DOI:** 10.1101/2020.12.18.20248457

**Authors:** Valeria Kebets, Pauline Favre, Josselin Houenou, Mircea Polosan, Jean-Michel Aubry, Dimitri Van De Ville, Camille Piguet

**Author notes:** **Corresponding author:** Valeria Kebets, University of Geneva, Campus Biotech, Chemin des Mines 9, 1202 Geneva, Switzerland.

## Abstract

**Background:** Emotion dysregulation is central to the development and maintenance of psychopathology, and is common across many psychiatric disorders. Neurobiological models of emotion dysregulation involve the fronto-limbic brain network, including in particular the amygdala and prefrontal cortex (PFC). Neural variability has recently been suggested as an index of cognitive flexibility. We hypothesized that within-subject neural variability in the fronto-limbic network would be related to inter-individual variation in emotion dysregulation in the context of low affective control.

**Methods:** In a multi-site cohort (*N* = 166, 93 females) of healthy individuals and individuals with emotional dysregulation (attention deficit/hyperactivity disorder (ADHD), bipolar disorder (BD), and borderline personality disorder (BPD)), we applied partial least squares (PLS), a multivariate data-driven technique, to derive latent components yielding maximal covariance between blood-oxygen level-dependent (BOLD) signal variability at rest and emotion dysregulation, as expressed by affective lability, depression and mania scores.

**Results:** PLS revealed one significant latent component (*r* = 0.62, *p* = 0.001), whereby greater emotion dysregulation was associated with increased neural variability in the amygdala, hippocampus, ventromedial, dorsomedial and dorsolateral PFC, insula and motor cortex, and decreased neural variability in occipital regions. This spatial pattern bears a striking resemblance to the fronto-limbic network, which is thought to subserve emotion regulation, and is impaired in individuals with ADHD, BD, and BPD.

**Conclusions:** Our work supports emotion dysregulation as a transdiagnostic dimension with neurobiological underpinnings that transcend diagnostic boundaries, and adds evidence to neural variability being a relevant proxy of neural efficiency.

## 1. Introduction

Emotion regulation allows individuals to modulate, manage or organize emotions in order to help them meet the demands of the environment and achieve their goals (Campos et al., 1989; Gross, 1998; Hilt et al., 2011), implicating various processes and systems (e.g., cognitive, behavioural, social, biological). In contrast, emotion dysregulation has been described as “a pattern of emotional experience and/or expression that interferes with appropriate goal-directed behavior” (Beauchaine, 2015). Emotion dysregulation is a central feature of psychopathology, and is key to both the development and maintenance of mood, personality and anxiety disorders, among others (Aldao et al., 2010; Sheppes et al., 2015; Weissman et al., 2019). Because it is both a risk factor for psychopathology in the general population, and is common across many forms of psychiatric disorders, a better understanding of the neurobiological underpinnings of emotion dysregulation may have important clinical applications, for instance in predicting or measuring the efficacy of a therapeutic intervention (Fowler et al., 2016; Sloan et al., 2017) in a transdiagnostic setting.

Emotion regulation is thought to rely on a fronto-limbic network, whereby the prefrontal cortex (PFC) exerts cognitive control over the amygdala, a subcortical structure that is central to emotion processing and salience perception (Adolphs, 2002; LeDoux, 1996; Ochsner et al., 2012). Unsurprisingly, alterations in this network are central to the pathophysiology of bipolar disorder (BD), borderline personality disorder (BPD), and attention deficit/hyperactivity disorder (ADHD), which are all characterized by emotion dysregulation (Chase and Phillips, 2016; Phillips and Swartz, 2014; Ruocco and Carcone, 2016; Schulze et al., 2016; Shaw et al., 2014; van Zutphen et al., 2015). Notably, the three disorders also share risk factors, such as childhood trauma and genetic overlap (Moukhtarian et al., 2018; Perroud et al., 2014; van Hulzen et al., 2017; Witt et al., 2017). This suggests that emotion dysregulation might have underlying neurobiological mechanisms that are shared across these disorders.

A measure that has received increasing attention in the past few years is neural variability, obtained by computing within-subject BOLD signal variability over the timecourse. First considered as neural “noise”, it has since been proposed as an index of local system dynamics (McIntosh et al., 2010). Indeed, a certain level of instability is thought to be required for the brain to flexibly explore different functional network configurations and adapt to various environmental demands (Deco et al., 2009; Ghosh et al., 2008; McIntosh et al., 2008). Neural variability has been shown to vary with age (Andrews-Hanna et al., 2007; Garrett et al., 2011, 2010; Guitart-Masip et al., 2016; Nomi et al., 2017; Samanez-Larkin et al., 2010), task performance (Armbruster-Genç et al., 2016; Garrett et al., 2011; Guitart-Masip et al., 2016; Raja Beharelle et al., 2012), but also symptom severity (Conio et al., 2019; Easson and McIntosh, 2019; Martino et al., 2016; Nomi et al., 2018). However, these relationships are often not linear (e.g., inverted U-shape in development and aging), and are task-, difficulty- and circuit-dependent (Armbruster-Genç et al., 2016; Garrett et al., 2014; Mišić et al., 2010). Nevertheless, this body of work demonstrates the functional relevance of neural variability, and that it can provide meaningful information that is complementary to mean-based measures.

Studies investigating neural variability in clinical populations have implicated neural circuits that are relevant for psychopathology. Indeed, brain signal variability in the medial PFC during rest was shown to correlate positively with increased ADHD symptoms and inattention in children with and without ADHD (Nomi et al., 2018). Furthermore, brain signal variability has been shown to vary with mood shifts. In patients with BD, opposing patterns of neural variability were found in the default and sensorimotor networks (SMN) between patients in the depressed and manic phases (Martino et al., 2016). This pattern mirrored the psychomotor behavior (i.e., acceleration/slowing), as well as the affective state (external/internal focus) that characterize the manic and depressive phases, respectively. Similarly, higher brain signal variability in the SMN was shown in individuals with a cyclothymic temperament compared to those with a depressive temperament in the general population (Conio et al., 2019). Interestingly, it was suggested that increased neuronal variability in specific circuits might facilitate local neuronal responses to incoming stimuli, and lead to over-excitation of specific behaviors/symptoms, e.g., psychomotor behavior, ruminations (Conio et al., 2019; Martino et al., 2016). However, to date, most studies looking at neural variability have relied on case-control comparisons, and few have tested for transnosographic, dimensional relationships.

In contrast to traditional case-control reports, a recent movement in psychiatry has advocated for a dimensional approach in the search for neurobiological markers of psychiatric symptoms. The NIMH’s Research Domain Criteria (RDoC) framework is one of the initiatives working towards developing a neurobiologically-based classification of mental disorders that integrates findings from behavioral science, neuroscience, and genetics (Cuthbert, 2014; Insel et al., 2010). Consequently, we favored a transdiagnostic approach in the present work by leveraging a multi-site cohort of healthy individuals and individuals suffering from conditions strongly associated with emotion dysregulation (ADHD, BD, BPD). We aimed to identify an emotion dysregulation dimension with associated patterns of blood-oxygen level-dependent (BOLD) variability, present in varying degrees among all individuals from our transdiagnostic cohort, which would suggest common neurobiological mechanisms that transcend diagnostic boundaries. We relied on partial least squares, a multivariate data-driven technique that extracts latent components by maximizing covariance between spatial patterns of neural variability and behavior (here, emotion dysregulation, as expressed by a combination of affective lability, depression and mania assessments). More specifically, we hypothesized that neural variability in the fronto-limbic circuit would be related to individual variation in emotion dysregulation.

## 2. Methods and Materials

### 2.1 Participants

Data for this study were collected from three sites (Geneva, Paris, Grenoble; see **Supplementary Figure S1** for participants’ inclusion and exclusion criteria). All participants gave their written informed consent. The research was conducted according to the principles of the Declaration of Helsinki and was approved by the University of Geneva research Ethics Committee (CER 13–081), the Paris CPP Ile de France IX Ethics Committee, and the Grenoble University Hospital Ethics Committee (n° 2011-A00425-36). Inclusion criteria for all participants were age between 18 and 55, no history of alcohol or drug abuse/dependence, no current or past cardiac or neurological disease. Exclusion criteria for all participants were a history of neurological disease or head trauma with loss of consciousness, any significant cerebral anatomic abnormality, and contraindications for MRI.

Patients with BD were recruited from the outpatient Mood Disorder Program of the Geneva University Hospital, from two university-affiliated participating centers (AP-HP, Henri Mondor Hospitals Créteil and Fernand Widal-Lariboisière Hospitals, Paris, France), and from the expert center for BD of Grenoble University Hospital. The clinical diagnosis was established using the DSM-IV-TR criteria by specialized psychiatrists and confirmed by the Mini-International Neuropsychiatric Interview (Sheehan et al., 1998), the Structured Clinical Interview for the DSM-IV (First et al., 2002), or the Diagnostic Interview for Genetic Studies (DIGS) (Nurnberger et al., 1994). Individuals were under stable medication for four weeks. Patients in Grenoble and Geneva were included in the study if they reported having been euthymic for at least one month prior to scanning and if they had a MADRS score < 15 and a YMRS score < 7. Patients in Paris were not in the acute phase of BD at the time of scanning.

BPD and ADHD patients were recruited from the outpatient Emotional Dysregulation Unit for BPD and ADHD patients of the Geneva University Hospital. BPD diagnosis was established with the SCID for DSM-IV Axis II Personality Disorders (First et al., 1997), and ADHD diagnosis with the Diagnostic Interview for ADHD in Adults (DIVA 2.0), by trained clinicians as part of the standard procedure of these specialized programs. Some patients were under psychotropic medication for comorbidities, as reported in **Table 1**. Participants were instructed not to take psychostimulants on the day of the study data acquisition.

**Table 1.** Demographic, imaging, and clinical profile of the sample used in the analyses (*N* = 166).

|  | **Bipolar**<br>**(N=75)** | **ADHD**<br>**(N=20)** | **Borderline**<br>**(N=21)** | **Controls**<br>**(N=65)** | **F/**<br>**chi<sup>2</sup>** | **p val** |
| --- | --- | --- | --- | --- | --- | --- |
| *Demographics* |  |  |  |  |  |  |
| Age, mean (SD) | 37.22 (11.61) | 24.00 (3.45) | 27.05 (4.67) | 35.06 (12.01) | 10.82 | **1.61E-06** |
| Sex (F/M) | 30 / 33 | 7 / 13 | 19 / 0 | 37 / 27 | 20.39 | **1.41E-04** |
| Education, mean (SD) <sup>a</sup> | 13.22 (2.70) | 16.00 (2.81) | 14.84 (3.18) | 12.67 (2.86) | 8.34 | **3.97E-05** |
| *Imaging* |  |  |  |  |  |  |
| Scanner (1/2/3/4) | 16 / 21 / 10 / 16 | 20 / 0 / 0 / 0 | 19 / 0 / 0 / 0 | 16 / 48 / 0 / 0 | 68.21 | **1.03E-14** |
| Framewise displacement, mean (SD) | 0.17 (0.09) | 0.14 (0.05) | 0.13 (0.04) | 0.17 (0.07) | 7.59 | **8.83E-05** |
| *Clinical* |  |  |  |  |  |  |
| ALS, mean (SD) | 1.04 (0.64) | 1.12 (0.48) | 1.80 (0.46) | 0.42 (0.40) | 39.44 | **3.44E-19** |
| MADRS, mean (SD) | 4.84 (4.51) | 3.55 (3.44) | 7.68 (3.45) | 1.23 (2.17) | 20.90 | **1.69E-11** |
| YMRS, mean (SD) | 1.68 (1.87) | 0.00 (0.00) | 1.47 (1.39) | 0.66 (1.39) | 8.77 | **2.00E-05** |
| *Disease severity* |  |  |  |  |  |  |
| Disease duration, mean (SD) <sup>b</sup> | 14.59 (9.34) | 6.80 (5.31) | 9.23 (5.39) | - | - | - |
| # Hospitalizations, mean (SD) <sup>c</sup> | 3.95 (3.24) | 0.05 (0.22) | 1.94 (2.54) | - | - | - |
| *Medication (by target) <sup>d</sup>* |  |  |  |  |  |  |
| Dopaminergic, No. (%) | 44 (70%) | 15 (75%) | 1 (5%) | 0 (0%) | - | - |
| Serotonergic, No. (%) | 46 (73%) | 1 (5%) | 2 (11%) | 0 (0%) | - | - |
| Glutamatergic, No. (%) | 42 (67%) | 0 (0%) | 0 (0%) | 0 (0%) | - | - |
| GABAergic, No. (%) | 36 (57%) | 0 (0%) | 0 (0%) | 0 (0%) | - | - |
| Norepinephrinergic, No. (%) | 30 (48%) | 15 (75%) | 0 (0%) | 0 (0%) | - | - |
| Lithium, No. (%) | 38 (60%) | 0 (0%) | 0 (0%) | 0 (0%) | - | - |
| No medication, No. (%) | 30 (48%) | 5 (25%) | 17 (89%) | 46 (72%) | - | - |
| Medication load, mean (SD) | 2.14 (1.51) | 0.85 (0.59) | 0.16 (0.50) | 0.31 (0.53) | - | - |

Control participants were recruited via local databases as well as through web advertisement and were matched with patients in terms of age, sex, level of education, and handedness. Exclusion criteria were past or present neurological or psychiatric disorders (Geneva, Grenoble), personal or family history of Axis I mood disorder, schizophrenia, or schizoaffective disorder (Paris), use of psychotropic medication, and contraindication for MRI. All participants underwent clinical assessment by trained raters using the DIGS (Nurnberger et al., 1994).

In total, 122 individuals with BD were recruited on all three sites, 93 healthy controls (HC) were recruited on two sites (i.e., Geneva and Paris), while 24 individuals with BPD and 21 individuals with ADHD were only recruited on one site (i.e., Geneva). We excluded 10 participants (8 BD, 1 BPD, 1 HC) for excessive in-scanner motion; 69 participants (39 BD, 2 BPD, 1 ADHD, 27 HC) because they had not completed the clinical measures of interest; 9 participants because they scored above 15 on the MADRS (7 BD, 2 BPD) and 6 because they scored above 7 on the YMRS (5 BD, 1 HC). The final sample thus comprised 166 participants, including 63 euthymic BD, 20 ADHD, 19 BPD, and 64 HC. The demographic, imaging, and clinical data of the final sample are shown in **Table 1**.

### 2.2 Clinical assessment

We used the Affective Lability Scales (ALS; (Harvey et al., 1989)), the Montgomery-Åsberg Depression Rating Scale (MADRS; (Montgomery and Åsberg, 1979)), and the Young Mania Rating Scale (YMRS; (Young et al., 1978)) to measure different facets of emotion dysregulation. The ALS specifically measures affective lability, which refers to the frequency, speed, and range of changes in affective states (Aas et al., 2015). The ALS is a 54-item self-reported questionnaire on which participants rate the tendency of their mood to shift between a “normal state” and different affects (depression, anger, anxiety and elation), as well as their tendency to experience shifts between elation and depression, and between anxiety and depression. The total score was obtained by averaging across the 6 subscales, i.e., anger, anxiety, anxiety/depression, depression, depression/elation, elation. The MADRS and YMRS are both clinician-rated scales that evaluate depressive and manic symptoms, respectively. The total score (sum across all items) was used for both scales.

### 2.3 Magnetic resonance imaging acquisition

Briefly, participants were scanned on 3T MRI scanners (see detailed MRI acquisition parameters in the **Supplementary Methods**). A resting state (RS) functional magnetic resonance imaging (fMRI) sequence, as well as an anatomical scan were acquired in all participants.

### 2.4 Rs fMRI preprocessing

The 10 first RS functional images were discarded to ensure signal equilibration, and the remaining images were preprocessed using SPM12 tools (http://fil.ion.ucl.ac.uk/spm/software/spm12). Functional images were first realigned, followed by co-registration of the mean functional image with the anatomical scan. Functional images were normalized to the MNI space with SPM12 “Segment”, resampled to 3mm isotropic voxels, and then spatially smoothed with a 6mm full-width-at-half-maximum Gaussian filter. The average signal within a mask of white matter (WM) and cerebrospinal fluid (CSF) were extracted using the Data Processing Assistant for Resting-State fMRI toolbox (Yan and Zang, 2010). The effects of WM, CSF and 6 motion parameters were regressed out from the time-course, and a bandpass filter (0.01 - 0.10 Hz) was applied. Motion scrubbing (Power et al., 2012) was applied to correct for motion artefacts; i.e., framewise displacement (FD) was calculated as the sum of the absolute values of the six realignment parameters, and scans with a FD higher than 0.5 mm, as well as one scan before and two scans after, were excluded from the analysis. Participants with a time-course containing less than 4mn of scanning were excluded (8 BD, 1 BPD, 1 HC).

### 2.5 DARTEL group template

A group template was generated with the Diffeomorphic Anatomical Registration Through Exponentiated Lie Algebra (DARTEL (Ashburner, 2007)) from the grey matter and white matter tissue segments of all the participants comprising the entire original sample (*N* = 250, see Figure S1). Participants’ T1 images were first segmented using the Computational Anatomy Toolbox (CAT12; http://www.neuro.uni-jena.de/cat/) “Segment Data” and the tissue segments were normalized to the tissue probability maps by means of an affine transformation. The group template was then normalized to the MNI space, and additional registration to the ICBM template was applied. Finally, the template was downsampled in order to match the dimensions of the functional images, and then binarized to include only voxels with a ≥ 50% grey matter probability.

Because of incomplete cerebellar coverage in 33 participants, we decided to exclude the cerebellum from the DARTEL template. To do so, we used a bilateral mask of the cerebellum as defined in Hammers atlas (Gousias et al., 2008; Hammers et al., 2003), a probabilistic anatomical atlas based on 83 manually-delineated regions drawn on MR images of 30 healthy adult subjects. In order to encompass the whole cerebellum, we first smoothed the mask with a 25mm FWHM Gaussian filter, then downsampled the mask to match the dimensions of the DARTEL template, and excluded the cerebellum mask from the DARTEL template.

### 2.6 BOLD signal variability

Voxel-wise BOLD signal variability was obtained for each participant by computing the standard deviation of each preprocessed time-course. This approach is equivalent to the frequency-domain computation of the amplitude of low-frequency fluctuations (ALFF) between 0.01 and 0.1 Hz (Garrett et al., 2013; Zöller et al., 2017), and is strongly correlated with mean-square successive differences (MSSD) (Garrett et al., 2011; Kielar et al., 2016). BOLD signal variability maps were constrained to the binarized DARTEL template (excluding the cerebellum), and were z-scored across all voxels included in the template (Zöller et al., 2017). Age, sex, scanner, and head motion (mean FD) were linearly regressed out from the imaging data prior to the PLS analysis using a general linear model on MATLAB.

### 2.7 Partial least squares analysis

We used partial least squares (PLS) analysis to identify BOLD variability spatial patterns related to emotion dysregulation across all participants. PLS is a multivariate data-driven statistical technique that aims to maximize covariance between two matrices (Krishnan et al., 2011; McIntosh and Lobaugh, 2004). The optimal relationship between the two data matrices is represented as latent components (LCs), which are weighted linear combinations of the original data that maximally covary with each other. A LC is characterized by a spatial pattern of neural variability and a behavioral pattern of affective lability, depression, and mania (imaging and behavioral saliences, respectively). By linearly projecting the imaging and behavioral measures of each participant onto their respective saliences, we obtain individual-specific brain and behavior scores, which reflect the participants’ imaging and behavioral contribution to each LC. Importantly, the PLS analysis was agnostic on the diagnostic group, so that transdiagnostic brain-behavior associations could be extracted.

The statistical significance of the LCs was assessed by constructing a null distribution of the singular values using permutation testing (1’000 permutations), whereby the behavioral data was permuted within each diagnostic group, so that latent components would not be driven by group differences. FDR correction (*q* < 0.05) was applied when assessing significance of the LCs. To determine which behavioral measures and voxels were driving the significant LC, we computed Pearson’s correlations between the original imaging data and brain scores, as well as between the original behavioral measures and behavioral scores (Courville and Thompson, 2001; Henson, 2002). A higher positive (or negative) correlation for a particular behavioral measure for a given LC indicates greater importance of the behavioral measure for the LC, while a higher positive (or negative) correlation for a particular imaging measure for a given LC indicates greater importance of that imaging value for the LC. We estimated confidence intervals for these correlations with a bootstrapping procedure that generated 1’000 samples with replacement from participants’ imaging and behavioral data, while accounting for diagnostic groups (i.e., bootstrap resampling was performed within each diagnostic group) in order to avoid spatial and behavioral patterns being driven by group differences, since our aim was to find transdiagnostic patterns of emotion dysregulation. Z-scores were computed by dividing each correlation coefficient by its bootstrap-estimated standard deviation, and were considered as strong contributors to LCs at absolute values >3, corresponding to a robustness at a confidence interval of approximately 99% (McIntosh and Lobaugh, 2004). See **Supplementary Methods** for more details.

### 2.8 Posthoc analyses

Two-sample t-tests were performed to test whether brain and behavioral scores for LC1 were different between participants from different diagnostic groups. Group differences in demographics, head motion, and clinical measures were tested using one-way analysis of variance (ANOVA, for continuous measures) or chi-squared tests (for categorical measures). We also tested if there were any significant associations between PLS brain (or behavioral) scores and disease severity, as well as medication use, using either Pearson’s correlations (for continuous measures), or t-tests (for binary measures). All posthoc analyses were corrected for multiple comparisons at a false discovery rate (FDR) of *q* < 0.05.

### 2.9 Control analyses

A number of control analyses were computed to assess the robustness of our results (detailed in the **Supplementary Methods**). Briefly, we used BOLD signal variability maps that included the cerebellum (*N* = 133); we accounted for education level (*N* = 138), early life trauma (*N* = 138), or disease severity (*N* = 55-82); we considered patients only (*N* = 102); and we considered only participants from the one site that included individuals from all four diagnostic groups to control for scan effects (*N* = 71).

### 2.10 Code availability

The code for the MRI preprocessing, BOLD signal variability extraction, as well as the PLS outputs can be found on <GITHUB_LINK> while the code for the PLS analysis is publicly available at https://github.com/danizoeller/myPLS.

## 3. Results

### 3.1 Neural variability correlates of emotion dysregulation

We applied PLS to whole-brain BOLD signal variability and a combination of clinical measures that characterize emotion dysregulation in 166 participants that were either healthy or had a diagnostic of ADHD, BD, or BPD. PLS revealed one significant latent component (LC1) that survived FDR correction (*q* < 0.05). LC1 revealed a significant association (*r* = 0.62, *p* = 0.001) between BOLD signal variability and emotion dysregulation (**Figure 1a**), accounting for 74% of the covariance between the two modalities.

**Figure 1.**
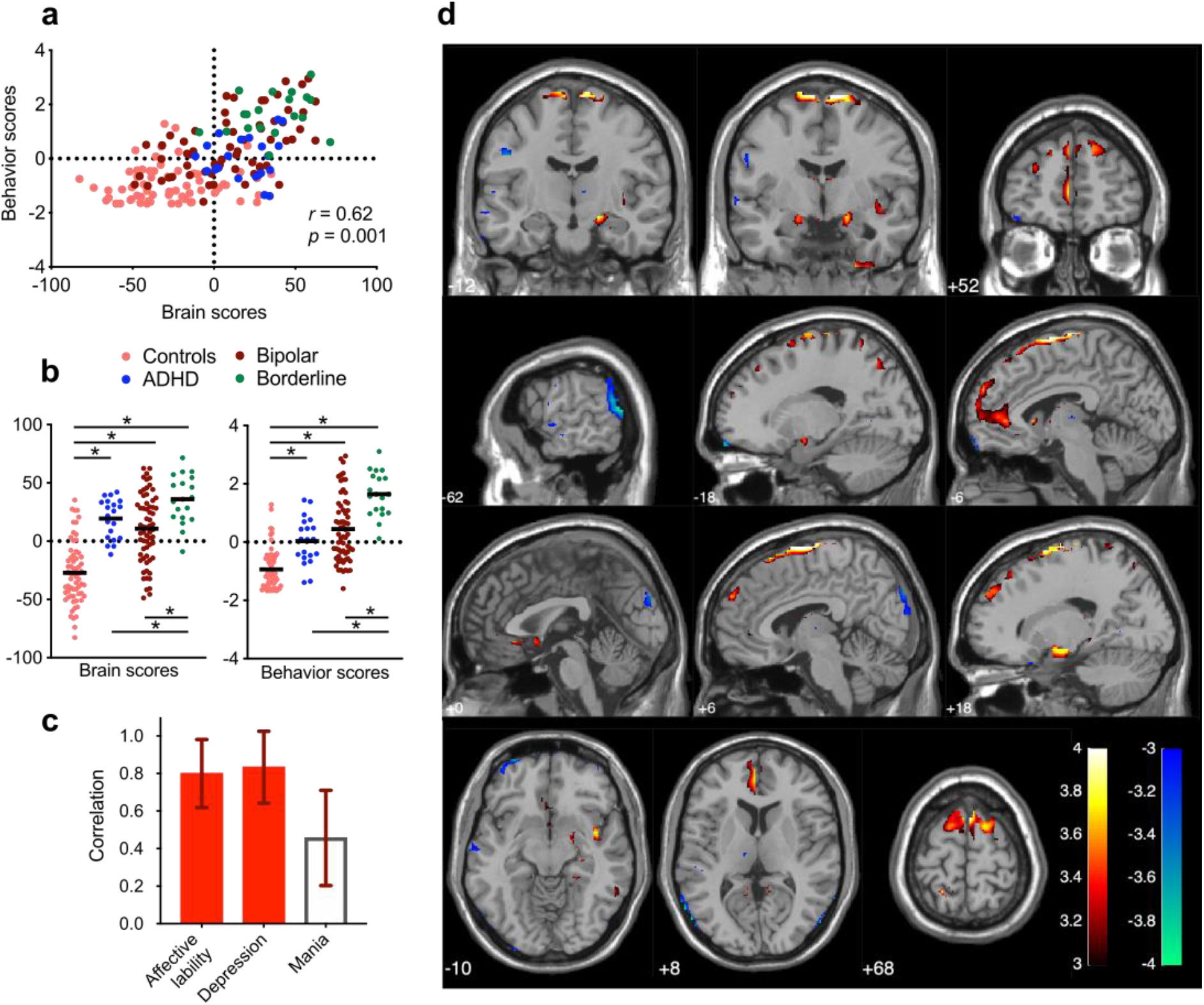
The first latent component (LC1) is characterized by high emotion dysregulation. (a) PLS correlation between individual brain and behavior scores for LC1. Each dot is a participant from any of the four diagnostic groups. (b) Group differences in brain and behavior scores for LC1. Bold lines are mean scores for each group, while asterisks indicate two-sample t-tests that have survived to FDR correction (*q* < 0.05). Controls have significantly lower brain and behavior scores compared to all patient groups. BPD patients have significantly higher brain and behavior scores compared to ADHD and BD patients. (c) Greater depression and affective lability characterize the behavioral pattern of LC1 (see **Supplementary Table S2**). Mania did not have a strong contribution to LC1 (*z* < 3). Loadings are Pearson’s correlations between participants’ original behavioral data and their behavior scores, and error bars indicate bootstrap-estimated standard deviations. (d) LC1 is characterized by increased BOLD signal variability in the left ventromedial PFC, bilateral dorsomedial PFC, subgenual ACC, bilateral amygdala, right hippocampus, right insula, and bilateral motor cortex, as well as decreased BOLD signal variability in occipital regions (see **Supplementary Table S3** for MNI coordinates of peaks of all significant clusters). Loadings are z-scores obtained from bootstrapping, thresholded at absolute values ≥ 3 (*p* < 0.01).

Healthy individuals had significantly lower brain and behavior scores compared to all patients groups (**Figure 1b**). Moreover, individuals with BPD had significantly higher brain and behavior scores compared to individuals with ADHD and BD. This dimension was therefore expressed more strongly by individuals with BPD, and less strongly by individuals with no psychiatric diagnosis, although there was a large overlap among individuals from all diagnostic groups (as shown on **Figure 1a**).

Higher loadings on LC1 were associated with greater affective lability and depression, whereas mania did not yield a strong contribution (**Figure 1c** and **Supplementary Table S2**). On the imaging side, LC1 was characterized by increased brain signal variability in the left ventromedial PFC, bilateral dorsomedial PFC, subgenual ACC, bilateral amygdala, right hippocampus, bilateral motor cortex, and right insula, as well as decreased variability in occipital regions (**Figure 1d** and **Supplementary Table S3** for the list of all reliable peaks and their MNI coordinates). This pattern recapitulates the emotion regulation network, which is known to be dysfunctional in ADHD, BD and BPD (Chase and Phillips, 2016; Phillips and Swartz, 2014; Ruocco and Carcone, 2016; Schulze et al., 2016; Shaw et al., 2014; van Zutphen et al., 2015).

Post-hoc associations between brain (or behavior scores) and disease severity, as well as medication use, can be found in **Table 2** and **Supplementary Table S4**. Medication targeting the dopaminergic system was associated with higher brain scores, while using medication affecting the serotonergic system was associated with higher behavioral scores. When categorizing medication use by medication class, the use of stimulants was also associated with higher brain scores.

**Table 2.** Post-hoc associations between participants’ brain (or behavioral) scores, disease severity, and medication use (categorized by the neurotransmitter target). Pearson correlations (for continuous measures) or t-tests (for categorical measures) were computed across all participants. Significant correlations or t-tests that survived FDR correction (*q* > 0.05) are indicated in bold. The same analysis was performed after classifying medication by medication class (see **Supplementary Table S4**).

|  | Brain scores |  | Behavior scores |  |
| --- | --- | --- | --- | --- |
|  | *r / t* | *p* | *r / t* | *p* |
| *Disease severity* |  |  |  |  |
| Disease duration | 0.11 | 0.334 | 0.01 | 0.958 |
| # Hospitalizations | -0.16 | 0.253 | 0.16 | 0.244 |
| *Medication use (by target)* |  |  |  |  |
| Dopaminergic | -2.40 | **0.018** | -2.10 | 0.037 |
| Serotonergic | 0.10 | 0.924 | -2.61 | **0.010** |
| Glutamatergic | 0.18 | 0.860 | 0.04 | 0.969 |
| GABAergic | 0.81 | 0.417 | -1.46 | 0.146 |
| Norepinephrinergic | -2.25 | 0.026 | -0.89 | 0.376 |
| Lithium | 1.87 | 0.064 | -0.73 | 0.467 |
| No medication | -0.99 | 0.325 | 0.74 | 0.458 |
| # Meds | -0.12 | 0.153 | 0.14 | 0.090 |

### 3.2 Control analyses

Control analyses whereby we i) used BOLD signal variability maps including the cerebellum; ii) accounted for education level or early life trauma; iii) considered patients only (i.e., excluded control participants); and iv) considered only participants from one site, all yielded saliences that were similar to the original brain and behavior saliences (see **Supplementary Table S5**), all showing the reliability of our findings. Detailed results are reported in the **Supplementary Results**.

## 4. Discussion

In this work, we aimed to identify spatial patterns of neural variability related to emotion dysregulation in a multi-site transdiagnostic cohort, using a multivariate data-driven approach. We found that emotion dysregulation was associated with a pattern of increased BOLD signal variability in the ventromedial PFC, dorsomedial and dorsolateral PFC, amygdala, hippocampus, insula and motor cortex, and decreased BOLD signal variability in occipital regions. Our findings are in line with emotion dysregulation being a dimensional construct that spans across individuals with affective disorders such as ADHD, BD and BPD, rather than being specific to any of these disorders. Importantly, the spatial pattern of brain signal variability associated with this dimension bears a compelling resemblance to the fronto-limbic circuit that is thought to subserve emotion regulation, and is impaired in ADHD, BD, and BPD. Our findings therefore add evidence to brain signal variability being a relevant proxy of neural efficiency, and support emotion dysregulation as a transdiagnostic dimension with neurobiological underpinnings that transcend diagnostic boundaries.

The patterns of brain signal variability associated with greater emotion dysregulation were mainly located in the fronto-limbic system, which plays a key role in emotional control/regulation (Ochsner et al., 2012; Ochsner and Gross, 2005). Critically, abnormalities in the fronto-limbic network are thought to underpin emotion dysregulation in pathophysiological models of BD and BPD (Chase and Phillips, 2016; Phillips and Swartz, 2014; Ruocco and Carcone, 2016; Schulze et al., 2016; van Zutphen et al., 2015). The suggested mechanism involves hyper-activation of limbic regions responsible for emotion generation – in particular, the amygdala, hippocampus, and ventral striatum-, coupled with hypo-activation of the PFC, which is responsible for cognitive control. This circuit has shown structural abnormalities in individuals with BD (Hanford et al., 2016; Hibar et al., 2018, 2016; Phillips and Swartz, 2014), BPD (Ruocco and Carcone, 2016; Schulze et al., 2016), but also ADHD (Hoogman et al., 2017), e.g., altered volumes of the amygdala and hippocampus, and cortical thinning of the PFC. In BPD patients, abnormal patterns of activity in the amygdala, hippocampus, vlPFC and dlPFC were shown during emotion processing (Ruocco et al., 2013; Ruocco and Carcone, 2016; Schulze et al., 2016), but also at rest, where the ACC, mPFC and dlPFC were found to be hyper-activated during resting state in BPD patients compared to control participants (Visintin et al., 2016). Furthermore, neural activation and connectivity of fronto-limbic regions, especially the ACC, amygdala, insula, and vlPFC, showed changes following psychotherapy aimed at improving emotion regulation in BPD patients (Marceau et al., 2018). Abnormal patterns of activity and connectivity of fronto-limbic regions have been reported in euthymic BD patients, especially involving the amygdala and the medial PFC at rest (Favre et al., 2014; Rey et al., 2016), and during emotion regulation tasks (Favre et al., 2015; Rey et al., 2014). Moreover, psychosocial intervention in individuals with BD or at risk for BD may induce functional and structural changes in these regions (Favre et al., 2016, 2013; Garrett et al., 2015). Emotion dysregulation is also prevalent in ADHD, and fronto-limbic alterations involving the amygdala, orbitofrontal cortex (OFC), ventral striatum and PFC have also been reported in this population (Shaw et al., 2014).

Previous studies of neural variability in ADHD (Depue et al., 2010; Mowinckel et al., 2017; Nomi et al., 2018; Sørensen et al., 2016), BD (Liu et al., 2012; Lu et al., 2014; Lui et al., 2015; Martino et al., 2016; Meda et al., 2015; Xu et al., 2014; Zhang et al., 2020), and BPD patients (Lei et al., 2017; Salvador et al., 2016) have failed to show any consistent pattern, although some have reported alterations in regions of the fronto-limbic network (mostly the PFC). In ADHD patients, increased BOLD signal variability in the dorsolateral PFC, inferior frontal and orbitofrontal cortex were found during a Stroop task (Depue et al., 2010), as well as increased brain signal variability in the ventromedial PFC during a vigilance task in adolescents with ADHD compared to controls (Sørensen et al., 2016). Moreover, greater MSSD in the dorsomedial PFC during rest was related to greater ADHD symptom severity, while greater MSSD in the ventromedial PFC was positively correlated with inattention across children with ADHD and typical developing children (Nomi et al., 2018). In BPD patients, increased ALFF was shown in the hippocampus (Salvador et al., 2016), while increased ALFF in the ventral PFC, dorsolateral PFC, and insula were found in euthymic BD patients (Xu et al., 2014), compared to controls. The fronto-limbic circuit also overlaps with the DMN, in particular the ventromedial PFC and hippocampus. We found increased neural variability of the ventromedial PFC to be associated with LC1, which was mostly driven by greater levels of depression. This partly corroborates a previous study contrasting neural variability patterns in the DMN and SMN, i.e., higher DMN/SMN ratio in the depressive phase of BD and the inverse pattern during mania, which were positively correlated with depression and mania scores, respectively (Martino et al., 2016). Therefore, our findings somewhat corroborate previous reports of altered neural variability in these clinical populations, but for the first time in a network directly associated with emotion regulation.

Our findings may be in apparent contrast to the prevailing view that brain signal variability facilitates neural flexibility by allowing fluid transitions between brain states via a stochastic resonance effect (Deco et al., 2009; Ghosh et al., 2008). However, while it appears to be beneficial to task performance, heightened neural variability was also shown to correlate with worse clinical symptoms in various conditions (Easson and McIntosh, 2019; Martino et al., 2016; Nomi et al., 2018). As neural variability is thought to increase the sensitivity to incoming stimuli, it is possible that heightened neural variability might lead to over-reactivity of specific neural circuits, as a maladaptive strategy to prepare for potentially relevant events, which instead supports the maintenance of emotion dysregulation in affective disorders. Consequently, our findings support the use of neural variability as a relevant proxy for dysfunctional emotional processing, which might be useful in tracking symptom severity and treatment efficacy (Dinstein et al., 2015).

The present study has several strengths including the use of a multivariate data-driven approach and the relatively large transdiagnostic cohort. Moreover, our approach aligns with recent initiatives such as the RDoC (Cuthbert, 2014; Insel et al., 2010), that promote neurobiologically-based approaches to investigate dimensions of (ab)normal functioning, which often transcend classical nosological categories. Computational techniques may help in this endeavor by deriving brain-behavior associations that could serve as potential phenotypes. Moreover, by quantifying their neurobiological correlates, data-driven machine learning approaches such as PLS could help refine future taxonomies for mental disorders.

However, this work also has some limitations. First, because of poor cerebellar coverage in a number of participants, we decided to exclude the cerebellum from our analyses, even though an increased number of reports implicate this structure in higher-order processes. Moreover, the diagnostic groups were not balanced across the three sites, which is a common shortcoming in multi-site studies with different protocols. Most patients were also using psychotropic medication at the time of scanning, which is known to modulate intrinsic brain activity metrics (Pereira-Sanchez et al., 2020). Our post-hoc analyses showed mild associations (*t* < 3) between LC1 and medication use. Finally, while PLS is a powerful technique for identifying brain-behavior associations, they remain correlational and do not claim to imply any causal relationship. Future studies are needed to further replicate these effects.

Despite some limitations, our findings have unveiled the neural variability correlates of emotion dysregulation in the fronto-limbic system, further improving our understanding of the pathogenesis of affective disorders. Importantly, emotion dysregulation is a transdiagnostic construct that has shown clinical utility as a therapeutic target, as demonstrated by a decrease in maladaptive emotion regulation strategy use and symptom severity (depression, anxiety, substance use, etc.), regardless of the treatment protocol, the construct of emotion regulation that was examined, and the targeted disorder (Sloan et al., 2017). Indeed, transdiagnostic protocols aimed at improving emotion regulation have been shown to provide rapid and significant improvement in individuals with various forms of severe mental illness (Fowler et al., 2016). In this context, our approach might also provide a robust way of tracking therapeutic effects of interventions aimed at enhancing emotion regulation.

## Data Availability

The data is available upon request.

## Acknowledgments

We wish to thank Gwladys Rey for her initial help in gathering a multisite database, Anne-Lise Küng and Eleonore Pham for their help in the recruitment of participants, and Alexandre Dayer for his inspirational guidance and enthusiasm for this project. He will stay in our hearts.

## References

Aas, M., Pedersen, G., Henry, C., Bjella, T., Bellivier, F., Leboyer, M., Kahn, J.-P., Cohen, R.F., Gard, S., Aminoff, S.R., Lagerberg, T.V., Andreassen, O.A., Melle, I., Etain, B., 2015. Psychometric properties of the Affective Lability Scale (54 and 18-item version) in patients with bipolar disorder, first-degree relatives, and healthy controls. J. Affect. Disord. 172, 375–380. https://doi.org/10.1016/j.jad.2014.10.028

Adolphs, R., 2002. Neural systems for recognizing emotion. Curr. Opin. Neurobiol. 12, 169–177. https://doi.org/10.1016/S0959-4388(02)00301-X

Aldao, A., Nolen-Hoeksema, S., Schweizer, S., 2010. Emotion-regulation strategies across psychopathology: A meta-analytic review. Clin. Psychol. Rev. 30, 217–237. https://doi.org/10.1016/j.cpr.2009.11.004

Andrews-Hanna, J.R., Snyder, A.Z., Vincent, J.L., Lustig, C., Head, D., Raichle, M.E., Buckner, R.L., 2007. Disruption of Large-Scale Brain Systems in Advanced Aging. Neuron 56, 924–935. https://doi.org/10.1016/j.neuron.2007.10.038

Armbruster-Genç, D.J.N., Ueltzhöffer, K., Fiebach, C.J., 2016. Brain signal variability differentially affects cognitive flexibility and cognitive stability. J. Neurosci. 36, 3978–3987. https://doi.org/10.1523/JNEUROSCI.2517-14.2016

Ashburner, J., 2007. A fast diffeomorphic image registration algorithm. NeuroImage 38, 95–113. https://doi.org/10.1016/j.neuroimage.2007.07.007

Beauchaine, T.P., 2015. Future Directions in Emotion Dysregulation and Youth Psychopathology. J. Clin. Child Adolesc. Psychol. Off. J. Soc. Clin. Child Adolesc. Psychol. Am. Psychol. Assoc. Div. 53 44, 875–896. https://doi.org/10.1080/15374416.2015.1038827

Campos, J.J., Campos, R.G., Barrett, K.C., 1989. Emergent themes in the study of emotional development and emotion regulation. Dev. Psychol. 25, 394–402. https://doi.org/10.1037/0012-1649.25.3.394

Chase, H.W., Phillips, M.L., 2016. Elucidating Neural Network Functional Connectivity Abnormalities in Bipolar Disorder: Toward a Harmonized Methodological Approach. Biol. Psychiatry Cogn. Neurosci. Neuroimaging 1, 288–298. https://doi.org/10.1016/j.bpsc.2015.12.006

Conio, B., Magioncalda, P., Martino, M., Tumati, S., Capobianco, L., Escelsior, A., Adavastro, G., Russo, D., Amore, M., Inglese, M., Northoff, G., 2019. Opposing patterns of neuronal variability in the sensorimotor network mediate cyclothymic and depressive temperaments. Hum. Brain Mapp. 40, 1344–1352. https://doi.org/10.1002/hbm.24453

Courville, T., Thompson, B., 2001. Use of structure coefficients in published multiple regression articles: β is not enough. Educ. Psychol. Meas. 61, 229–248.

Cuthbert, B.N., 2014. The RDoC framework: facilitating transition from ICD/DSM to dimensional approaches that integrate neuroscience and psychopathology. World Psychiatry 13, 28–35. https://doi.org/10.1002/wps.20087

Deco, G., Jirsa, V., McIntosh, A.R., Sporns, O., Kötter, R., 2009. Key role of coupling, delay, and noise in resting brain fluctuations. Proc. Natl. Acad. Sci. 106, 10302–10307. https://doi.org/10.1073/pnas.0901831106

Depue, B.E., Burgess, G.C., Willcutt, E.G., Bidwell, L.C., Ruzic, L., Banich, M.T., 2010. Symptom-correlated brain regions in young adults with combined-type ADHD: Their organization, variability, and relation to behavioral performance. Psychiatry Res. Neuroimaging 182, 96–102. https://doi.org/10.1016/j.pscychresns.2009.11.011

Dinstein, I., Heeger, D.J., Behrmann, M., 2015. Neural variability: friend or foe? Trends Cogn. Sci. 19, 322–328. https://doi.org/10.1016/j.tics.2015.04.005

Easson, A.K., McIntosh, A.R., 2019. BOLD signal variability and complexity in children and adolescents with and without autism spectrum disorder. Dev. Cogn. Neurosci. 36, 100630. https://doi.org/10.1016/j.dcn.2019.100630

Favre, P., Baciu, M., Pichat, C., Bougerol, T., Polosan, M., 2014. fMRI evidence for abnormal resting-state functional connectivity in euthymic bipolar patients. J. Affect. Disord. 165, 182–189. https://doi.org/10.1016/j.jad.2014.04.054

Favre, P., Baciu, M., Pichat, C., De Pourtalès, M.-A., Fredembach, B., Garçon, S., Bougerol, T., Polosan, M., 2013. Modulation of fronto-limbic activity by the psychoeducation in euthymic bipolar patients. A functional MRI study. Psychiatry Res. Neuroimaging 214, 285–295. https://doi.org/10.1016/j.pscychresns.2013.07.007

Favre, P., Houenou, J., Baciu, M., Pichat, C., Poupon, C., Bougerol, T., Polosan, M., 2016. White matter plasticity induced by Psychoeducation in bipolar patients: a controlled diffusion tensor imaging study. Psychother. Psychosom. 85, 58–60.

Favre, P., Polosan, M., Pichat, C., Bougerol, T., Baciu, M., 2015. Cerebral Correlates of Abnormal Emotion Conflict Processing in Euthymic Bipolar Patients: A Functional MRI Study. PloS One 10, e0134961. https://doi.org/10.1371/journal.pone.0134961

First, M.B., Gibbon, M., Spitzer, R.L., Benjamin, L.S., 1997. User’s guide for the structured clinical interview for DSM-IV axis II personality disorders: SCID-II. American Psychiatric Press, Washington, DC.

First, M.B., Spitzer, R.L., Gibbon, M., Williams, J.B.W., 2002. Structured Clinical Interview for DSM-IV-TR Axis I Disorders, Research version, Patient edition (SCID-I/P).

Fowler, J.C., Clapp, J.D., Madan, A., Allen, J.G., Oldham, J.M., Frueh, B.C., 2016. Emotion dysregulation as a cross-cutting target for inpatient psychiatric intervention. J. Affect. Disord. 206, 224–231. https://doi.org/10.1016/j.jad.2016.07.043

Garrett, A.S., Miklowitz, D.J., Howe, M.E., Singh, M.K., Acquaye, T.K., Hawkey, C.G., Glover, G.H., Reiss, A.L., Chang, K.D., 2015. Changes in brain activation following psychotherapy for youth with mood dysregulation at familial risk for bipolar disorder. Prog. Neuropsychopharmacol. Biol. Psychiatry 56, 215–220. https://doi.org/10.1016/j.pnpbp.2014.09.007

Garrett, D.D., Kovacevic, N., McIntosh, A.R., Grady, C.L., 2011. The importance of being variable. J. Neurosci. 31, 4496–4503. https://doi.org/10.1523/JNEUROSCI.5641-10.2011

Garrett, D.D., Kovacevic, N., McIntosh, A.R., Grady, C.L., 2010. Blood oxygen level-dependent signal variability is more than just noise. J. Neurosci. 30, 4914–4921. https://doi.org/10.1523/JNEUROSCI.5166-09.2010

Garrett, D.D., McIntosh, A.R., Grady, C.L., 2014. Brain Signal Variability is Parametrically Modifiable. Cereb. Cortex 24, 2931–2940. https://doi.org/10.1093/cercor/bht150

Garrett, D.D., Samanez-Larkin, G.R., MacDonald, S.W.S., Lindenberger, U., McIntosh, A.R., Grady, C.L., 2013. Moment-to-moment brain signal variability: a next frontier in human brain mapping? Neurosci. Biobehav. Rev. 37, 610–624. https://doi.org/10.1016/j.neubiorev.2013.02.015

Ghosh, A., Rho, Y., McIntosh, A.R., Kötter, R., Jirsa, V.K., 2008. Noise during rest enables the exploration of the brain’s dynamic repertoire. PLoS Comput. Biol. 4, e1000196. https://doi.org/10.1371/journal.pcbi.1000196

Gousias, I.S., Rueckert, D., Heckemann, R.A., Dyet, L.E., Boardman, J.P., Edwards, A.D., Hammers, A., 2008. Automatic segmentation of brain MRIs of 2-year-olds into 83 regions of interest. NeuroImage 40, 672–684. https://doi.org/10.1016/j.neuroimage.2007.11.034

Gross, J.J., 1998. The Emerging Field of Emotion Regulation: An Integrative Review. Rev. Gen. Psychol. 2, 271–299. https://doi.org/10.1037/1089-2680.2.3.271

Guitart-Masip, M., Salami, A., Garrett, D., Rieckmann, A., Lindenberger, U., Bäckman, L., 2016. BOLD Variability is Related to Dopaminergic Neurotransmission and Cognitive Aging. Cereb. Cortex 26, 2074–2083. https://doi.org/10.1093/cercor/bhv029

Hammers, A., Allom, R., Koepp, M.J., Free, S.L., Myers, R., Lemieux, L., Mitchell, T.N., Brooks, D.J., Duncan, J.S., 2003. Three-dimensional maximum probability atlas of the human brain, with particular reference to the temporal lobe. Hum. Brain Mapp. 19, 224–247. https://doi.org/10.1002/hbm.10123

Hanford, L.C., Nazarov, A., Hall, G.B., Sassi, R.B., 2016. Cortical thickness in bipolar disorder: a systematic review. Bipolar Disord. 18, 4–18. https://doi.org/10.1111/bdi.12362

Harvey, P.D., Greenberg, B.R., Serper, M.R., 1989. The affective lability scales: development, reliability, and validity. J. Clin. Psychol. 45, 786–793. https://doi.org/10.1002/1097-4679(198909)45:5<786::aid-jclp2270450515>3.0.co;2-p

Henson, R.K., 2002. The Logic and Interpretation of Structure Coefficients in Multivariate General Linear Model Analyses.

Hibar, D.P., Westlye, L.T., Doan, N.T., Jahanshad, N., Cheung, J.W., Ching, C.R.K., Versace, A., Bilderbeck, A.C., Uhlmann, A., Mwangi, B., Krämer, B., Overs, B., Hartberg, C.B., Abé, C., Dima, D., Grotegerd, D., Sprooten, E., Bøen, E., Jimenez, E., Howells, F.M., Delvecchio, G., Temmingh, H., Starke, J., Almeida, J.R.C., Goikolea, J.M., Houenou, J., Beard, L.M., Rauer, L., Abramovic, L., Bonnin, M., Ponteduro, M.F., Keil, M., Rive, M.M., Yao, N., Yalin, N., Najt, P., Rosa, P.G., Redlich, R., Trost, S., Hagenaars, S., Fears, S.C., Alonso-Lana, S., van Erp, T.G.M., Nickson, T., Chaim-Avancini, T.M., Meier, T.B., Elvsåshagen, T., Haukvik, U.K., Lee, W.H., Schene, A.H., Lloyd, A.J., Young, A.H., Nugent, A., Dale, A.M., Pfennig, A., McIntosh, A.M., Lafer, B., Baune, B.T., Ekman, C.J., Zarate, C.A., Bearden, C.E., Henry, C., Simhandl, C., McDonald, C., Bourne, C., Stein, D.J., Wolf, D.H., Cannon, D.M., Glahn, D.C., Veltman, D.J., Pomarol-Clotet, E., Vieta, E., Canales-Rodriguez, E.J., Nery, F.G., Duran, F.L.S., Busatto, G.F., Roberts, G., Pearlson, G.D., Goodwin, G.M., Kugel, H., Whalley, H.C., Ruhe, H.G., Soares, J.C., Fullerton, J.M., Rybakowski, J.K., Savitz, J., Chaim, K.T., Fatjó-Vilas, M., Soeiro-de-Souza, M.G., Boks, M.P., Zanetti, M.V., Otaduy, M.C.G., Schaufelberger, M.S., Alda, M., Ingvar, M., Phillips, M.L., Kempton, M.J., Bauer, M., Landén, M., Lawrence, N.S., van Haren, N.E.M., Horn, N.R., Freimer, N.B., Gruber, O., Schofield, P.R., Mitchell, P.B., Kahn, R.S., Lenroot, R., Machado-Vieira, R., Ophoff, R.A., Sarró, S., Frangou, S., Satterthwaite, T.D., Hajek, T., Dannlowski, U., Malt, U.F., Arolt, V., Gattaz, W.F., Drevets, W.C., Caseras, X., Agartz, I., Thompson, P.M., Andreassen, O.A., 2018. Cortical abnormalities in bipolar disorder: an MRI analysis of 6503 individuals from the ENIGMA Bipolar Disorder Working Group. Mol. Psychiatry 23, 932–942. https://doi.org/10.1038/mp.2017.73

Hibar, D.P., Westlye, L.T., van Erp, T.G.M., Rasmussen, J., Leonardo, C.D., Faskowitz, J., Haukvik, U.K., Hartberg, C.B., Doan, N.T., Agartz, I., Dale, A.M., Gruber, O., Krämer, B., Trost, S., Liberg, B., Abé, C., Ekman, C.J., Ingvar, M., Landén, M., Fears, S.C., Freimer, N.B., Bearden, C.E., Sprooten, E., Glahn, D.C., Pearlson, G.D., Emsell, L., Kenney, J., Scanlon, C., McDonald, C., Cannon, D.M., Almeida, J., Versace, A., Caseras, X., Lawrence, N.S., Phillips, M.L., Dima, D., Delvecchio, G., Frangou, S., Satterthwaite, T.D., Wolf, D., Houenou, J., Henry, C., Malt, U.F., Bøen, E., Elvsåshagen, T., Young, A.H., Lloyd, A.J., Goodwin, G.M., Mackay, C.E., Bourne, C., Bilderbeck, A., Abramovic, L., Boks, M.P., van Haren, N.E.M., Ophoff, R.A., Kahn, R.S., Bauer, M., Pfennig, A., Alda, M., Hajek, T., Mwangi, B., Soares, J.C., Nickson, T., Dimitrova, R., Sussmann, J.E., Hagenaars, S., Whalley, H.C., McIntosh, A.M., Thompson, P.M., Andreassen, O.A., 2016. Subcortical volumetric abnormalities in bipolar disorder. Mol. Psychiatry 21, 1710–1716. https://doi.org/10.1038/mp.2015.227

Hilt, L.M., Hanson, J.L., Pollack, S.D., 2011. Emotion dysregulation, in: Encyclopedia of Adolescence. Elsevier, New York, pp. 160–169.

Hoogman, M., Bralten, J., Hibar, D.P., Mennes, M., Zwiers, M.P., Schweren, L.S.J., van Hulzen, K.J.E., Medland, S.E., Shumskaya, E., Jahanshad, N., Zeeuw, P. de, Szekely, E., Sudre, G., Wolfers, T., Onnink, A.M.H., Dammers, J.T., Mostert, J.C., Vives-Gilabert, Y., Kohls, G., Oberwelland, E., Seitz, J., Schulte-Rüther, M., Ambrosino, S., Doyle, A.E., Høvik, M.F., Dramsdahl, M., Tamm, L., van Erp, T.G.M., Dale, A., Schork, A., Conzelmann, A., Zierhut, K., Baur, R., McCarthy, H., Yoncheva, Y.N., Cubillo, A., Chantiluke, K., Mehta, M.A., Paloyelis, Y., Hohmann, S., Baumeister, S., Bramati, I., Mattos, P., Tovar-Moll, F., Douglas, P., Banaschewski, T., Brandeis, D., Kuntsi, J., Asherson, P., Rubia, K., Kelly, C., Martino, A.D., Milham, M.P., Castellanos, F.X., Frodl, T., Zentis, M., Lesch, K.-P., Reif, A., Pauli, P., Jernigan, T.L., Haavik, J., Plessen, K.J., Lundervold, A.J., Hugdahl, K., Seidman, L.J., Biederman, J., Rommelse, N., Heslenfeld, D.J., Hartman, C.A., Hoekstra, P.J., Oosterlaan, J., Polier, G. von, Konrad, K., Vilarroya, O., Ramos-Quiroga, J.A., Soliva, J.C., Durston, S., Buitelaar, J.K., Faraone, S.V., Shaw, P., Thompson, P.M., Franke, B., 2017. Subcortical brain volume differences in participants with attention deficit hyperactivity disorder in children and adults: a cross-sectional mega-analysis. Lancet Psychiatry 4, 310–319. https://doi.org/10.1016/S2215-0366(17)30049-4

Insel, T., Cuthbert, B., Garvey, M., Heinssen, R., Pine, D.S., Quinn, K., Sanislow, C., Wang, P., 2010. Research domain criteria (RDoC): toward a new classification framework for research on mental disorders. Am. J. Psychiatry 167, 748–751. https://doi.org/10.1176/appi.ajp.2010.09091379

Kielar, A., Deschamps, T., Chu, R.K.O., Jokel, R., Khatamian, Y.B., Chen, J.J., Meltzer, J.A., 2016. Identifying Dysfunctional Cortex: Dissociable Effects of Stroke and Aging on Resting State Dynamics in MEG and fMRI. Front. Aging Neurosci. 8. https://doi.org/10.3389/fnagi.2016.00040

Krishnan, A., Williams, L.J., McIntosh, A.R., Abdi, H., 2011. Partial Least Squares (PLS) methods for neuroimaging: a tutorial and review. NeuroImage 56, 455–475. https://doi.org/10.1016/j.neuroimage.2010.07.034

LeDoux, J.E., 1996. The emotional brain: The mysterious underpinnings of emotional life, Simon&Schuster. ed. New York.

Lei, X., Zhong, M., Liu, Y., Jin, X., Zhou, Q., Xi, C., Tan, C., Zhu, X., Yao, S., Yi, J., 2017. A resting-state fMRI study in borderline personality disorder combining amplitude of low frequency fluctuation, regional homogeneity and seed based functional connectivity. J. Affect. Disord. 218, 299–305. https://doi.org/10.1016/j.jad.2017.04.067

Liu, C.-H., Ma, X., Wu, X., Li, F., Zhang, Y., Zhou, F.-C., Wang, Y.-J., Tie, C.-L., Zhou, Z., Zhang, D., Dong, J., Yao, L., Wang, C.-Y., 2012. Resting-state abnormal baseline brain activity in unipolar and bipolar depression. Neurosci. Lett. 516, 202–206. https://doi.org/10.1016/j.neulet.2012.03.083

Lu, D., Jiao, Q., Zhong, Y., Gao, W., Xiao, Q., Liu, X., Lin, X., Cheng, W., Luo, L., Xu, C., Lu, G., Su, L., 2014. Altered baseline brain activity in children with bipolar disorder during mania state: a resting-state study. Neuropsychiatr. Dis. Treat. 10, 317–323. https://doi.org/10.2147/NDT.S54663

Lui, S., Yao, L., Xiao, Y., Keedy, S.K., Reilly, J.L., Keefe, R.S., Tamminga, C.A., Keshavan, M.S., Pearlson, G.D., Gong, Q., Sweeney, J.A., 2015. Resting-state brain function in schizophrenia and psychotic bipolar probands and their first-degree relatives. Psychol. Med. 45, 97–108. https://doi.org/10.1017/S003329171400110X

Marceau, E.M., Meuldijk, D., Townsend, M.L., Solowij, N., Grenyer, B.F.S., 2018. Biomarker correlates of psychotherapy outcomes in borderline personality disorder: A systematic review. Neurosci. Biobehav. Rev. 94, 166–178. https://doi.org/10.1016/j.neubiorev.2018.09.001

Martino, M., Magioncalda, P., Huang, Z., Conio, B., Piaggio, N., Duncan, N.W., Rocchi, G., Escelsior, A., Marozzi, V., Wolff, A., 2016. Contrasting variability patterns in the default mode and sensorimotor networks balance in bipolar depression and mania. Proc. Natl. Acad. Sci. 113, 4824–4829.

McIntosh, A.R., Kovacevic, N., Itier, R.J., 2008. Increased brain signal variability accompanies lower behavioral variability in development. PLoS Comput. Biol. 4, e1000106. https://doi.org/10.1371/journal.pcbi.1000106

McIntosh, A.R., Kovacevic, N., Lippe, S., Garrett, D., Grady, C., Jirsa, V., 2010. The development of a noisy brain. Arch. Ital. Biol. 148, 323–337.

McIntosh, A.R., Lobaugh, N.J., 2004. Partial least squares analysis of neuroimaging data: applications and advances. NeuroImage 23 Suppl 1, S250–263. https://doi.org/10.1016/j.neuroimage.2004.07.020

Meda, S.A., Wang, Z., Ivleva, E.I., Poudyal, G., Keshavan, M.S., Tamminga, C.A., Sweeney, J.A., Clementz, B.A., Schretlen, D.J., Calhoun, V.D., Lui, S., Damaraju, E., Pearlson, G.D., 2015. Frequency-Specific Neural Signatures of Spontaneous Low-Frequency Resting State Fluctuations in Psychosis: Evidence From Bipolar-Schizophrenia Network on Intermediate Phenotypes (B-SNIP) Consortium. Schizophr. Bull. 41, 1336–1348. https://doi.org/10.1093/schbul/sbv064

Mišić, B., Mills, T., Taylor, M.J., McIntosh, A.R., 2010. Brain Noise Is Task Dependent and Region Specific. J. Neurophysiol. 104, 2667–2676. https://doi.org/10.1152/jn.00648.2010

Montgomery, S.A., Åsberg, M., 1979. A New Depression Scale Designed to be Sensitive to Change. Br. J. Psychiatry 134, 382–389. https://doi.org/10.1192/bjp.134.4.382

Moukhtarian, T.R., Mintah, R.S., Moran, P., Asherson, P., 2018. Emotion dysregulation in attention-deficit/hyperactivity disorder and borderline personality disorder. Borderline Personal. Disord. Emot. Dysregulation 5, 9. https://doi.org/10.1186/s40479-018-0086-8

Mowinckel, A.M., Alnæs, D., Pedersen, M.L., Ziegler, S., Fredriksen, M., Kaufmann, T., Sonuga-Barke, E., Endestad, T., Westlye, L.T., Biele, G., 2017. Increased default-mode variability is related to reduced task-performance and is evident in adults with ADHD. NeuroImage Clin. 16, 369–382. https://doi.org/10.1016/j.nicl.2017.03.008

Nomi, J.S., Bolt, T.S., Ezie, C.E.C., Uddin, L.Q., Heller, A.S., 2017. Moment-to-moment BOLD signal variability reflects regional changes in neural flexibility across the lifespan. J. Neurosci. 37, 5539–5548. https://doi.org/10.1523/JNEUROSCI.3408-16.2017

Nomi, J.S., Schettini, E., Voorhies, W., Bolt, T.S., Heller, A.S., Uddin, L.Q., 2018. Resting-State Brain Signal Variability in Prefrontal Cortex Is Associated With ADHD Symptom Severity in Children. Front. Hum. Neurosci. 12, 90. https://doi.org/10.3389/fnhum.2018.00090

Nurnberger, J.I., Blehar, M.C., Kaufmann, C.A., York-Cooler, C., Simpson, S.G., Harkavy-Friedman, J., Severe, J.B., Malaspina, D., Reich, T., 1994. Diagnostic interview for genetic studies. Rationale, unique features, and training. NIMH Genetics Initiative. Arch. Gen. Psychiatry 51, 849–859; discussion 863-864. https://doi.org/10.1001/archpsyc.1994.03950110009002

Ochsner, K.N., Gross, J.J., 2005. The cognitive control of emotion. Trends Cogn. Sci. 9, 242–249. https://doi.org/10.1016/j.tics.2005.03.010

Ochsner, K.N., Silvers, J.A., Buhle, J.T., 2012. Functional imaging studies of emotion regulation: a synthetic review and evolving model of the cognitive control of emotion. Ann. N. Y. Acad. Sci. 1251, E1–E24. https://doi.org/10.1111/j.1749-6632.2012.06751.x

Pereira-Sanchez, V., Franco, A.R., Vieira, D., de Castro-Manglano, P., Soutullo, C., Milham, M.P., Castellanos, F.X., 2020. Systematic Review: Medication Effects on Brain Intrinsic Functional Connectivity in Patients With Attention-Deficit/Hyperactivity Disorder. J. Am. Acad. Child Adolesc. Psychiatry. https://doi.org/10.1016/j.jaac.2020.10.013

Perroud, N., Cordera, P., Zimmermann, J., Michalopoulos, G., Bancila, V., Prada, P., Dayer, A., Aubry, J.-M., 2014. Comorbidity between attention deficit hyperactivity disorder (ADHD) and bipolar disorder in a specialized mood disorders outpatient clinic. J. Affect. Disord. 168, 161–166. https://doi.org/10.1016/j.jad.2014.06.053

Phillips, M.L., Swartz, H.A., 2014. A Critical Appraisal of Neuroimaging Studies of Bipolar Disorder: Toward a New Conceptualization of Underlying Neural Circuitry and a Road Map for Future Research. Am. J. Psychiatry 171, 829–843. https://doi.org/10.1176/appi.ajp.2014.13081008

Power, J.D., Barnes, K.A., Snyder, A.Z., Schlaggar, B.L., Petersen, S.E., 2012. Spurious but systematic correlations in functional connectivity MRI networks arise from subject motion. NeuroImage 59, 2142–2154. https://doi.org/10.1016/j.neuroimage.2011.10.018

Raja Beharelle, A., Kovačević, N., McIntosh, A.R., Levine, B., 2012. Brain signal variability relates to stability of behavior after recovery from diffuse brain injury. NeuroImage 60, 1528–1537. https://doi.org/10.1016/j.neuroimage.2012.01.037

Rey, G., Desseilles, M., Favre, S., Dayer, A., Piguet, C., Aubry, J.-M., Vuilleumier, P., 2014. Modulation of brain response to emotional conflict as a function of current mood in bipolar disorder: preliminary findings from a follow-up state-based fMRI study. Psychiatry Res. Neuroimaging 223, 84–93.

Rey, G., Piguet, C., Benders, A., Favre, S., Eickhoff, S.B., Aubry, J.-M., Vuilleumier, P., 2016. Resting-state functional connectivity of emotion regulation networks in euthymic and non-euthymic bipolar disorder patients. Eur. Psychiatry 34, 56–63. https://doi.org/10.1016/j.eurpsy.2015.12.005

Ruocco, A.C., Amirthavasagam, S., Choi-Kain, L.W., McMain, S.F., 2013. Neural Correlates of Negative Emotionality in Borderline Personality Disorder: An Activation-Likelihood-Estimation Meta-Analysis. Biol. Psychiatry, Risk Mechanisms for Bipolar Disorder 73, 153–160. https://doi.org/10.1016/j.biopsych.2012.07.014

Ruocco, A.C., Carcone, D., 2016. A Neurobiological Model of Borderline Personality Disorder: Systematic and Integrative Review. Harv. Rev. Psychiatry 24, 311–329. https://doi.org/10.1097/HRP.0000000000000123

Salvador, R., Vega, D., Pascual, J.C., Marco, J., Canales-Rodríguez, E.J., Aguilar, S., Anguera, M., Soto, A., Ribas, J., Soler, J., Maristany, T., Rodríguez-Fornells, A., Pomarol-Clotet, E., 2016. Converging Medial Frontal Resting State and Diffusion-Based Abnormalities in Borderline Personality Disorder. Biol. Psychiatry, Borderline Personality Disorder: Mechanisms of Emotion Dysregulation 79, 107–116. https://doi.org/10.1016/j.biopsych.2014.08.026

Samanez-Larkin, G.R., Kuhnen, C.M., Yoo, D.J., Knutson, B., 2010. Variability in nucleus accumbens activity mediates age-related suboptimal financial risk taking. J. Neurosci. 30, 1426–1434. https://doi.org/10.1523/JNEUROSCI.4902-09.2010

Schulze, L., Schmahl, C., Niedtfeld, I., 2016. Neural Correlates of Disturbed Emotion Processing in Borderline Personality Disorder: A Multimodal Meta-Analysis. Biol. Psychiatry, Borderline Personality Disorder: Mechanisms of Emotion Dysregulation 79, 97–106. https://doi.org/10.1016/j.biopsych.2015.03.027

Shaw, P., Stringaris, A., Nigg, J., Leibenluft, E., 2014. Emotion dysregulation in attention deficit hyperactivity disorder. Am. J. Psychiatry 171, 276–293. https://doi.org/10.1176/appi.ajp.2013.13070966

Sheppes, G., Suri, G., Gross, J.J., 2015. Emotion regulation and psychopathology. Annu. Rev. Clin. Psychol. 11, 379–405. https://doi.org/10.1146/annurev-clinpsy-032814-112739

Sloan, E., Hall, K., Moulding, R., Bryce, S., Mildred, H., Staiger, P.K., 2017. Emotion regulation as a transdiagnostic treatment construct across anxiety, depression, substance, eating and borderline personality disorders: A systematic review. Clin. Psychol. Rev. 57, 141–163. https://doi.org/10.1016/j.cpr.2017.09.002

Sørensen, L., Eichele, T., van Wageningen, H., Plessen, K.J., Stevens, M.C., 2016. Amplitude variability over trials in hemodynamic responses in adolescents with ADHD: The role of the anterior default mode network and the non-specific role of the striatum. NeuroImage Clin. 12, 397–404. https://doi.org/10.1016/j.nicl.2016.08.007

van Hulzen, K.J.E., Scholz, C.J., Franke, B., Ripke, S., Klein, M., McQuillin, A., Sonuga-Barke, E.J., Kelsoe, J.R., Landén, M., Andreassen, O.A., Lesch, K.-P., Weber, H., Faraone, S.V., Arias-Vasquez, A., Reif, A., 2017. Genetic Overlap Between Attention-Deficit/Hyperactivity Disorder and Bipolar Disorder: Evidence From Genome-wide Association Study Meta-analysis. Biol. Psychiatry, Attention-Deficit/Hyperactivity Disorder: Predictors of Treatment Response and Comorbidities 82, 634–641. https://doi.org/10.1016/j.biopsych.2016.08.040

van Zutphen, L., Siep, N., Jacob, G.A., Goebel, R., Arntz, A., 2015. Emotional sensitivity, emotion regulation and impulsivity in borderline personality disorder: a critical review of fMRI studies. Neurosci. Biobehav. Rev. 51, 64–76. https://doi.org/10.1016/j.neubiorev.2015.01.001

Visintin, E., De Panfilis, C., Amore, M., Balestrieri, M., Wolf, R.C., Sambataro, F., 2016. Mapping the brain correlates of borderline personality disorder: A functional neuroimaging meta-analysis of resting state studies. J. Affect. Disord. 204, 262–269. https://doi.org/10.1016/j.jad.2016.07.025

Weissman, D.G., Bitran, D., Miller, A.B., Schaefer, J.D., Sheridan, M.A., McLaughlin, K.A., 2019. Difficulties with emotion regulation as a transdiagnostic mechanism linking child maltreatment with the emergence of psychopathology. Dev. Psychopathol. 31, 899–915. https://doi.org/10.1017/S0954579419000348

Witt, S.H., Streit, F., Jungkunz, M., Frank, J., Awasthi, S., Reinbold, C.S., Treutlein, J., Degenhardt, F., Forstner, A.J., Heilmann-Heimbach, S., Dietl, L., Schwarze, C.E., Schendel, D., Strohmaier, J., Abdellaoui, A., Adolfsson, R., Air, T.M., Akil, H., Alda, M., Alliey-Rodriguez, N., Andreassen, O.A., Babadjanova, G., Bass, N.J., Bauer, M., Baune, B.T., Bellivier, F., Bergen, S., Bethell, A., Biernacka, J.M., Blackwood, D.H.R., Boks, M.P., Boomsma, D.I., Børglum, A.D., Borrmann-Hassenbach, M., Brennan, P., Budde, M., Buttenschøn, H.N., Byrne, E.M., Cervantes, P., Clarke, T.- K., Craddock, N., Cruceanu, C., Curtis, D., Czerski, P.M., Dannlowski, U., Davis, T., de Geus, E.J.C., Di Florio, A., Djurovic, S., Domenici, E., Edenberg, H.J., Etain, B., Fischer, S.B., Forty, L., Fraser, C., Frye, M.A., Fullerton, J.M., Gade, K., Gershon, E.S., Giegling, I., Gordon, S.D., Gordon-Smith, K., Grabe, H.J., Green, E.K., Greenwood, T.A., Grigoroiu-Serbanescu, M., Guzman-Parra, J., Hall, L.S., Hamshere, M., Hauser, J., Hautzinger, M., Heilbronner, U., Herms, S., Hitturlingappa, S., Hoffmann, P., Holmans, P., Hottenga, J.-J., Jamain, S., Jones, I., Jones, L.A., Juréus, A., Kahn, R.S., Kammerer-Ciernioch, J., Kirov, G., Kittel-Schneider, S., Kloiber, S., Knott, S.V., Kogevinas, M., Landén, M., Leber, M., Leboyer, M., Li, Q.S., Lissowska, J., Lucae, S., Martin, N.G., Mayoral-Cleries, F., McElroy, S.L., McIntosh, A.M., McKay, J.D., McQuillin, A., Medland, S.E., Middeldorp, C.M., Milaneschi, Y., Mitchell, P.B., Montgomery, G.W., Morken, G., Mors, O., Mühleisen, T.W., Müller-Myhsok, B., Myers, R.M., Nievergelt, C.M., Nurnberger, J.I., O’Donovan, M.C., Loohuis, L.M.O., Ophoff, R., Oruc, L., Owen, M.J., Paciga, S.A., Penninx, B.W.J.H., Perry, A., Pfennig, A., Potash, J.B., Preisig, M., Reif, A., Rivas, F., Rouleau, G.A., Schofield, P.R., Schulze, T.G., Schwarz, M., Scott, L., Sinnamon, G.C.B., Stahl, E.A., Strauss, J., Turecki, G., Van der Auwera, S., Vedder, H., Vincent, J.B., Willemsen, G., Witt, C.C., Wray, N.R., Xi, H.S., Tadic, A., Dahmen, N., Schott, B.H., Cichon, S., Nöthen, M.M., Ripke, S., Mobascher, A., Rujescu, D., Lieb, K., Roepke, S., Schmahl, C., Bohus, M., Rietschel, M., 2017. Genome-wide association study of borderline personality disorder reveals genetic overlap with bipolar disorder, major depression and schizophrenia. Transl. Psychiatry 7, e1155–e1155. https://doi.org/10.1038/tp.2017.115

Xu, K., Liu, H., Li, H., Tang, Y., Womer, F., Jiang, X., Chen, K., Zhou, Y., Jiang, W., Luo, X., Fan, G., Wang, F., 2014. Amplitude of low-frequency fluctuations in bipolar disorder: A resting state fMRI study. J. Affect. Disord. 152–154, 237–242. https://doi.org/10.1016/j.jad.2013.09.017

Yan, C., Zang, Y., 2010. DPARSF: a MATLAB toolbox for “pipeline” data analysis of resting-state fMRI. Front. Syst. Neurosci. 4. https://doi.org/10.3389/fnsys.2010.00013

Young, R.C., Biggs, J.T., Ziegler, V.E., Meyer, D.A., 1978. A rating scale for mania: reliability, validity and sensitivity. Br. J. Psychiatry J. Ment. Sci. 133, 429–435.

Zhang, Z., Bo, Q., Li, F., Zhao, L., Wang, Y., Liu, R., Chen, X., Wang, C., Zhou, Y., 2020. Increased ALFF and functional connectivity of the right striatum in bipolar disorder patients. Prog. Neuropsychopharmacol. Biol. Psychiatry 110140. https://doi.org/10.1016/j.pnpbp.2020.110140

Zohar, J., Nutt, D.J., Kupfer, D.J., Moller, H.-J., Yamawaki, S., Spedding, M., Stahl, S.M., 2014. A proposal for an updated neuropsychopharmacological nomenclature. Eur. Neuropsychopharmacol. J. Eur. Coll. Neuropsychopharmacol. 24, 1005–1014. https://doi.org/10.1016/j.euroneuro.2013.08.004

Zohar, J., Stahl, S., Moller, H.-J., Blier, P., Kupfer, D., Yamawaki, S., Uchida, H., Spedding, M., Goodwin, G.M., Nutt, D., 2015. A review of the current nomenclature for psychotropic agents and an introduction to the Neuroscience-based Nomenclature. Eur. Neuropsychopharmacol. J. Eur. Coll. Neuropsychopharmacol. 25, 2318–2325. https://doi.org/10.1016/j.euroneuro.2015.08.019

Zöller, D., Schaer, M., Scariati, E., Padula, M.C., Eliez, S., Van De Ville, D., 2017. Disentangling resting-state BOLD variability and PCC functional connectivity in 22q11.2 deletion syndrome. NeuroImage 149, 85–97. https://doi.org/10.1016/j.neuroimage.2017.01.064

