## Supplemental Information for "Fronto-limbic neural variability as a transdiagnostic correlate of emotion dysregulation"

#### Supplementary Methods

##### *MRI acquisition parameters*

Geneva (Site 1)

MRI data were acquired at the University of Geneva on a 3T Siemens MAGNETOM TrioTim scanner with a 32-channel head coil. A resting state functional magnetic resonance imaging (rs-fMRI) sequence as well as an anatomical T1-weighted scan were acquired. Whole brain functional images were collected using a BOLD-weighted EPI sequence (TR/TE = 2100/30 ms; flip angle = 80 degrees; PAT factor = 2; FOV = 205 mm; matrix size = 64 x 64 pixels). Thirty-six transversal slices were acquired sequentially with a 3.2 mm thickness and an interslice gap of 3.84 mm, yielding a voxel size of 3.2 x 3.2 x 3.2 mm. 250 volumes were acquired for a total duration of 8 minutes 45 seconds. Subjects were instructed to close their eyes and let their thoughts wander, and not to fall asleep. A high-resolution whole brain anatomical scan was acquired with a T1-weighted 3D sequence (MPRAGE; TR/TI/TE = 1900/900/2.27 ms; flip angle = 9 degrees; voxel dimensions = 1.0 mm isotropic).

Paris (Site 2)

MRI data were collected at Neurospin (Commissariat à l'Energie Atomique, Saclay, France) using a 3T Siemens Magnetom TrioTim with a 12-channel head coil. The rs-fMRI images were collected using the following acquisition parameters: EPI sequence; TR/TE = 2000/27 ms; flip angle = 81 degrees, FOV = 192 mm, voxel size = 3 x 3 x 3 mm. 360 volumes were acquired for a total duration of 12 minutes. Participants were instructed to close their eyes and try not to think about anything specific, without falling asleep. A high-resolution T1-weighted anatomical scan was also acquired (MPRAGE; TR/TI/TE = 2300/900/2.98 ms; flip angle = 9 degrees; voxel dimensions = 1 x 1 x 1.1 mm).

Grenoble (Sites 3 and 4)

Imaging data were acquired using two whole-body 3T MR scanners (Bruker MedSpec S300 and Achieva 3.0 TX Philips) at the Grenoble MRI facility IRMaGE with a similar fMRI acquisition sequence. Sixteen and 28 patients with BD were scanned on the first and second scanner, respectively. Functional images were acquired using the following acquisition parameters for both scanners: interleaved EPI sequence, T2*-weighted; TR/TE = 2500/30 ms; flip angle = 77 degrees; FOV = 216 mm; matrix size = 72 x 72 pixels). Thirty-nine slices were acquired transversally on the Bruker scanner, with a 3.5 mm thickness, and an interslice gap of 3 mm, yielding a voxel size of 3 x 3 x 3.5 mm. Thirty-seven slices were acquired transversally on the Philips scanner with a 3 mm thickness, an interslice gap of 3 mm, and a voxel size of 3 x 3 x 3.75 mm. volumes were acquired on both scanners for a total duration of 6 minutes. Subjects were asked to stay awake with their eyes open and to not think about anything in particular. A high-resolution whole brain anatomical scan was acquired with a T1-weighted 3D sequence (TR/TI/TE = 1900/900/2.27 ms; flip angle = 9 degrees; voxel dimensions = 0.8 mm isotropic; acquisition matrix 280 x 320 x 220 pixels). A 3D Modified Driven Equilibrium Fourier Transform (MDEFT) and a turbo field echo (TFE) sequence were used for acquisitions using the Bruker and the Philips scanners, respectively.

##### *Medication use*

Medication was categorized by the affected neurotransmitter system, according to the Neuroscience-based Nomenclature (NbN-2, (Zohar et al. 2014, 2015), https://nbn2r.com/). Targeted neurotransmitters included the dopaminergic, GABAergic, glutamatergic, lithium, norepinephrinergic, and serotonergic. Posthoc analyses involving the use of “other” medication were not tested, because these medications were very heterogeneous and not necessarily psychotropic. In a control analysis, we also classified medication by medication class (e.g., antidepressants, mood stabilizers, etc.). The list of all medications used by participants, and their categorization into targeted neurotransmitter(s) and medication class can be found in **Supplementary Table S1**.

##### *Partial least squares analysis*

Partial least squares (PLS) is a multivariate data-driven statistical technique that aims to extract latent components (also called latent variables), representing optimal brain-behavior associations (McIntosh and Lobaugh 2004; Krishnan et al. 2011). While it is mechanistically similar to canonical correlation analysis (CCA), PLS has the advantage of not suffering from rank deficiency when the number of features is higher than the number of sample, or in case of high autocorrelation (e.g., spatial autocorrelation or correlation between behavioral variables), which makes it ideally suited for neuroimaging data (McIntosh and Mišić 2013).

BOLD signal variability maps are stored in $X$ (participants × voxels), while the clinical data are stored in $Y$ (participants × behavioral measures). After z-scoring both matrices across all participants, we computed a cross-covariance matrix $R$:

$$R = Y^T X$$

followed by singular value decomposition of $R$, which provides the reconstruction of $R$ by matrices $U$, $S$, and $V$:

$$R = USV^T$$

The singular vectors $V$ and $U$ are called the *brain* and *behavior saliences* (akin to loadings in principal components analysis), while $S$ is a diagonal matrix containing the singular values. Next, we computed $L_X$ and $L_Y$ by projecting $X$ and $Y$ onto their respective saliences $V$ and $U$**:**

$$L_X = XV$$

$$L_Y = YU$$

The matrices obtained (one for the imaging data and one for the behavior data, per LC) are called *brain* and *behavior scores* (akin to the scores in principal components analysis) and reflect the participants' imaging and behavioral contribution to each LC.

The covariance explained by each LC is estimated by dividing the squared singular value by the sum of all squared singular values, and the LCs are ordered by the amount of covariance they account for.

The contribution of the original variables to the LC relied on *structure coefficients*, i.e., Pearson's correlations between $X$ and $L_X$, as well as between $Y$ and $L_Y$ (Courville and Thompson 2001; Henson 2002). Structure coefficients are commonly used in factor analysis, multiple regression analysis and CCA, as they help to define the structure of the synthetic variables, and thereby reflect the direct contribution of a predictor to the predictor criterion (independently of other predictors), which can be critical when predictors are highly correlated between each other (i.e., in presence of multicollinearity (Sherry and Henson 2005)). Notably, structure coefficients were high correlated to saliences in our data ($r$ = 0.90, $r$ = 1 with brain and behavior saliences of LC1, respectively).

##### *Control analyses*

A number of control analyses were computed to assess the robustness of our results. To begin with, we tested if there were any significant associations between PLS brain (or behavioral) scores and disease severity, as well as medication use. These posthoc analyses were corrected for multiple comparisons at a false discovery rate (FDR) of $q < 0.05$.

We also ran a number of control PLS analyses (detailed below), whereby we examined the similarity (i.e., Pearson correlation) between the saliences (i.e., brain and behavioral) obtained in the original PLS model and the saliences obtained in the control analyses. First, since recent reports have implicated the cerebellum in emotion and cognitive processes (Green et al. 2007; Buckner 2013; Moberget et al. 2018), the PLS analysis was re-computed after including the cerebellum in BOLD signal variability maps ($N = 133$), after excluding participants with incomplete cerebellar coverage. Second, we re-ran the analysis in participants with available education level data ($N = 138$), after adding it to the other regressors. Furthermore, there is substantial evidence showing that early life trauma impacts the clinical expression of mood disorders (Aas et al. 2014); therefore, we re-computed the analysis in participants who were administered the Childhood Trauma Questionnaire ($N = 138$) after adding its total score to the regressors. In order to test whether the LCs were driven by group differences between controls and patients, the PLS analysis was re-computed only in patients. Because scanner can crucially impact imaging measures, we re-ran our analysis by including only participants from Geneva, which was the only site that included individuals from all four diagnostic groups.

Finally, in order to account for disease severity, we regressed disease duration and number of hospitalizations from the imaging data before re-computing PLS.

### Supplementary Results

#### *Control analyses*

Including the cerebellum in BOLD signal variability maps (*N* = 133) yielded very similar patterns (Pearson *r* = 0.94, *r* = 1 with the original brain and behavior saliences of LC1, respectively). After accounting for education level (*N* = 138), the brain-behavior association remained significant (*p* = 0.006), and very similar to the original results (*r* = 0.65, *r* = 0.90 with the original brain and behavior saliences, respectively). When accounting for early life trauma (*N* = 138), results were moderately similar to the original analysis (*r* = -0.54, *r* = 0.40 with the original brain and behavior saliences, respectively). When considering only patients (i.e., excluding control participants from the analysis), results were very similar to the original analysis (*r* = 0.65, *r* = 1 with the original brain and behavior saliences, respectively). To account for scanner effects, we re-ran permutation testing (i.e., a null distribution of the singular values) accounting for scanner, and found that LC1 remained significant (*p* = 0.021). We also computed another control analysis considering participants from only one site (Geneva; *N* = 71) as it was the only site that included individuals from all four diagnostic groups, and we found moderate to very high similarity with the original results (*r* = -0.50, *r* = 0.98 with brain and behavior saliences, respectively).

When accounting for disease severity (i.e., disease duration, number of hospitalizations), we found that LC1 was no longer significant; this may be due to the low sample size, as many patients were missing these information (*N* between 55 and 82).

**Table S1. Medication sorted by targeted neurotransmitter system(s) and medication class.** Categorization by targeted neurotransmitter was based on the Neuroscience-based Nomenclature (NbN-2 (Zohar et al. 2014, 2015); https://nbn2r.com/). Possible neurotransmitter targets were the acetylcholinergic, dopaminergic, epinephrinergic, GABAergic, glutamatergic, histaminic, lithium, norepinephrinergic, and serotonergic systems, as well as "other" (which included medication not targeting any neurotransmitter system). Medication targeting the acetylcholinergic, epinephrinergic, and histaminic systems were merged with "other" as these categories had too few participants (≤2) on their own. Medication classes comprised typical and atypical antipsychotics, antidepressants, mood stabilizers, sedatives/hypnotics/anxiolytics (SHA), stimulants, and "other".

| Molecule / Medication | Targeted neurotransmitter(s) | Medication class |
|---|---|---|
| Alimemazine / Theralene | Dopamine, Histamine | Typical antipsychotic |
| Almotriptan / Almogran | Serotonin | Antidepressant |
| Amisulpride / Solian | Dopamine | Atypical antipsychotic |
| Amlodipine / Amlor | Other | Other |
| Aripiprazole / Abilify | Dopamine, Serotonin | Atypical antipsychotic |
| Atomoxetine / Strattera | Norepinephrine | Stimulant |
| Biopros | Other | Other |
| Bupropion / Wellbutrin | Dopamine, Norepinephrine | Antidepressant |
| Buspirone | Serotonin | SHA |
| Carbamazepine / Tegretol | Glutamate | Mood stabilizer |
| Chlorpromazine / Largactil, Prazine | Dopamine, Serotonin | Typical antipsychotic |
| Citalopram / Seropram | Serotonin | Antidepressant |
| Clonazepam / Rivotril | GABA | SHA |
| Clorazepate / Potassium clorazepate | GABA | Other |
| Cyamemazine / Tercian | Dopamine, Serotonin | Typical antipsychotic |
| Dexmethylphenidate / Focalin | Dopamine, Norepinephrine | Stimulant |
| Diazepam / Valium | GABA | SHA |
| Eschscholtia | Other | Other |
| Escitalopram / Cipralex, Seroplex | Serotonin | Antidepressant |
| Estrogen / various birth control | Other | Other |
| Ezetimibe / Inegy | Other | Other |
| Fenofibrate | Other | Other |
| Fluoxetine / Prozac | Serotonin | Antidepressant |
| Flurazepam / Dalmadorm | GABA | SHA |
| Gabapentin / Neurontin | Glutamate | Other |
| Gestodene / various birth control | Other | Other |
| Iron, ferrous sulfate / Tardyferon | Other | Other |
| Lamotrigine / Lamictal | Glutamate | Mood stabilizer |
| Levomepromazine | Dopamine, Serotonin | Typical antipsychotic |
| Levonorgestrel / various birth control | Other | Other |
| Levothyroxine sodium | Other | Other |
| Lisdexamfetamine / Elvanse | Dopamine, Norepinephrine | Stimulant |
| Lithium / Lithiofor, Priadel, Teralithe | Lithium | Mood stabilizer |
| Lorazepam / Temesta | GABA | SHA |
| Loxapine | Dopamine, Serotonin | Typical antipsychotic |
| Melatonin / Circadin | Other | Other |

| | | |
|---|---|---|
| Methylphenidate / Ritaline, Concerta | Dopamine, Norepinephrine | Stimulant |
| Mirtazapine | Serotonin, Norepinephrine | Antidepressant |
| Nicotine / Nicopass, Nicorette | Other | Other |
| Nux vomica | Other | Other |
| Olanzapine / Zyprexa | Dopamine, Serotonin | Atypical antipsychotic |
| Oxazepam / Anxiolit | GABA | SHA |
| Oxcarbazepine / Trileptal | Glutamate | Mood stabilizer |
| Paroxetine / Deroxat | Serotonin | Antidepressant |
| Saccharomyces boulardii / Perenterol | Other | Other |
| Prazepam / Lysanxia | GABA | SHA |
| Progestin / various birth control | Other | Other |
| Progestogen / various birth control | Other | Other |
| Quetiapine / Seroquel, Xeroquel | Dopamine, Serotonin | Atypical antipsychotic |
| Rabeprazole / Pariet | Other | Other |
| Risperidone / Risperdal, Risperdaloro | Dopamine, Serotonin, Norepinephrine | Atypical antipsychotic |
| Salmeterol / other drugs for obstructive airway diseases | Epinephrine | Other |
| Sertraline / Zoloft | Serotonin | Antidepressant |
| Simvastatin / Inegy | Other | Other |
| Topiramate / Topamax | Glutamate, GABA | Mood stabilizer |
| Tropatepine / Lepticur | Acetylcholine | Other |
| Valproic acid, valproate / Depakin, Depakote, Orfiril | Glutamate | Mood stabilizer |
| Valpromide / Depamide | Glutamate | Mood stabilizer |
| Venlafaxine / Effexor | Serotonin, Norepinephrine | Antidepressant |
| Verapamil | Other | Other |
| Zolmitriptan / Zomig | Serotonin | Other |
| Zolpidem / Stillnox | GABA | SHA |
| Zopiclone / Imovane | GABA | SHA |

**Table S2. Contribution of each behavioral measure to LC1.** Pearson's correlations between participants' behavioral measures and their behavioral scores, along with their bootstrap-estimated standard deviations (SD) are shown. Absolute *Z* scores above 3 were considered reliable ($p > 0.01$) and are indicated in bold.

| Behavior variable | Correlation (SD) | Z-score |
|---|---|---|
| Affective lability (ALS) | 0.89 (0.18) | **4.40** |
| Depression (MADRS) | 0.83 (0.19) | **4.35** |
| Mania (YMRS) | 0.46 (0.25) | 1.80 |

**Table S3. Anatomical regions and MNI coordinates of BOLD signal variability clusters that reliably contributed to LC1** (absolute z-scores ≥ 3). Anatomical labels were determined using the Anatomy toolbox (v.3) in SPM12, which applies probabilistic algorithms to determine the cytoarchitectonic labeling of MNI coordinates (Eickhoff et al. 2005, 2006, 2007).

| Anatomical region | Cluster size | MNI coordinates | | | Z-score |
|---|---|---|---|---|---|
| | | x | y | z | |
| *Positive clusters* | | | | | |
| L Superior parietal lobule / Postcentral gyrus | 326 | -12 | -7 | 77 | 3.99 |
| R Hippocampus / Parahippocampal gyrus | 247 | 21 | -16 | -13 | 3.37 |
| R Superior frontal gyrus / Precentral gyrus | 220 | 12 | -1 | 74 | 3.70 |
| L Frontal pole / Paracingulate gyrus | 208 | -3 | 56 | 5 | 3.74 |
| R Superior frontal gyrus / Postcentral gyrus | 126 | 15 | -34 | 80 | 3.51 |
| R Frontal pole / Superior frontal gyrus | 124 | 18 | 56 | 35 | 3.38 |
| L Frontal pole / Superior frontal gyrus | 92 | -21 | 44 | 38 | 3.25 |
| R Insular cortex / Planum polare | 83 | 36 | -13 | 5 | 3.35 |
| L Anterior cingulate gyrus / Paracingulate gyrus | 67 | -3 | -4 | 41 | 3.13 |
| R Temporal pole / Frontal orbital cortex | 62 | 45 | 17 | -16 | 2.97 |
| L Superior frontal gyrus / Paracingulate gyrus | 39 | -3 | 50 | 35 | 2.80 |
| R Middle temporal gyrus / Inferior temporal gyrus | 36 | 60 | -52 | -7 | 2.94 |
| R Inferior temporal gyrus / Temporal occipital fusiform cortex | 35 | 51 | -49 | -19 | 3.49 |
| L Insular cortex / Planum polare | 30 | -36 | 8 | -16 | 2.90 |
| L Superior lateral occipital cortex | 28 | -21 | -70 | 44 | 3.08 |
| R Superior lateral occipital cortex | 26 | 27 | -67 | 47 | 3.19 |
| R Temporal fusiform cortex / Inferior temporal gyrus | 22 | 30 | -1 | -49 | 2.72 |
| L Temporal pole | 20 | -51 | 11 | -13 | 2.72 |
| *Negative clusters* | | | | | |
| L Inferior lateral occipital cortex / Occipital pole | 659 | -60 | -67 | 20 | 3.91 |
| L Frontal pole / Frontal medial cortex | 245 | -30 | 65 | -10 | 3.59 |
| R Frontal pole | 165 | 42 | 56 | -13 | 3.74 |
| L Postcentral gyrus / Precentral gyrus | 165 | -48 | -13 | 32 | 3.81 |
| R Cuneal cortex / Occipital pole | 162 | 3 | -88 | 29 | 3.21 |
| R Angular gyrus / Supramarginal gyrus | 147 | 60 | -61 | 29 | 3.67 |
| L Middle temporal gyrus | 133 | -54 | -13 | -16 | 3.33 |
| R Postcentral gyrus / Precentral gyrus | 89 | 69 | -7 | 17 | 3.56 |
| L Planum temporale / Heschl's gyrus | 63 | -54 | -19 | 5 | 3.27 |
| L Putamen / Frontal orbital cortex | 60 | -27 | 11 | -7 | 2.78 |
| L Inferior temporal gyrus | 58 | -54 | -37 | -28 | 3.83 |
| R Occipital pole / Inferior lateral occipital cortex | 55 | 36 | -94 | -7 | 3.02 |
| L Frontal orbital cortex / Frontal operculum cortex | 44 | -30 | 26 | -7 | 3.19 |
| R Frontal orbital cortex / Subcallosal cortex | 39 | 9 | 17 | -25 | 3.44 |

| | | | | | |
|---|---|---|---|---|---|
| L Frontal orbital cortex / Subcallosal cortex | 35 | -9 | 14 | -25 | 2.96 |
| R Inferior lateral occipital cortex | 35 | 54 | -76 | 5 | 3.31 |
| L Middle frontal gyrus / Inferior frontal gyrus, pars opercularis | 33 | -39 | 11 | 26 | 3.05 |
| L Middle temporal gyrus | 23 | -54 | -49 | 2 | 3.20 |
| R Inferior temporal gyrus | 20 | 54 | -28 | -31 | 2.86 |

**Table S4.** Post-hoc associations between participations' brain (or behavioral) scores and medication use (categorized by medication class). T-tests were used to compare brain (or behavioral scores) between participants that used one type of medication compared to participants that didn't use that type of medication. Significant t-tests that survived FDR correction ($q$ > 0.05) are indicated in bold.

| | Brain scores | | Behavior scores | |
|---|---|---|---|---|
| | *t* | *p* | *t* | *p* |
| *Medication use (by class)* | | | | |
| Typical antipsychotics | 1.38 | 0.170 | -1.69 | 0.093 |
| Atypical antipsychotics | -1.63 | 0.106 | -1.85 | 0.066 |
| Antidepressants | -0.12 | 0.902 | -2.08 | 0.039 |
| Mood stabilizers | 1.10 | 0.273 | -0.88 | 0.383 |
| Sedates/Hypnotics/Anxiolytics | 1.00 | 0.321 | -0.78 | 0.434 |
| Stimulants | -2.94 | **0.004** | -0.90 | 0.370 |

**Table S5.** Similarity (Pearson correlation) between the saliences of each control analysis and the original saliences.

| | *Including cerebellum* | *Regressing education* | *Regressing early life trauma* | *Patients only* | *Geneva only* |
|---|---|---|---|---|---|
| Correlations with original behavior saliences | 1.00 | 0.90 | 0.40 | 1.00 | 0.98 |
| Correlations with original imaging saliences | 0.94 | 0.65 | -0.54 | 0.65 | -0.50 |

**Figure S1.** Flowchart illustrating the composition of the original sample and final sample, and the number of subjects excluded based on different criteria. *Abbreviations: ADHD = attention deficit/hyperactivity disorder; BD = bipolar disorder; BPD = borderline personality disorder; HC= healthy controls.*

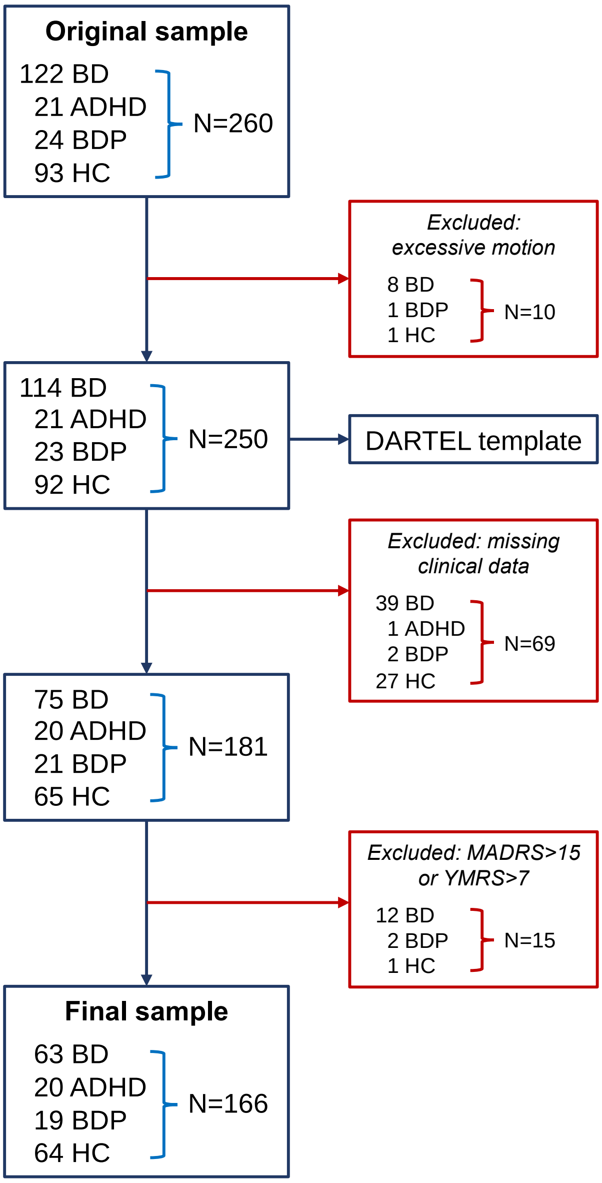

#### Supplementary References

Aas M, Aminoff SR, Vik Lagerberg T, Etain B, Agartz I, Andreassen OA, et al. Affective lability in patients with bipolar disorders is associated with high levels of childhood trauma. Psychiatry Res. 2014 Aug 15;218(1–2):252–5.

Buckner RL. The Cerebellum and Cognitive Function: 25 Years of Insight from Anatomy and Neuroimaging. Neuron. 2013 Oct;80(3):807–15.

Courville T, Thompson B. Use of structure coefficients in published multiple regression articles: β is not enough. Educ Psychol Meas. 2001;61(2):229–248.

Eickhoff SB, Heim S, Zilles K, Amunts K. Testing anatomically specified hypotheses in functional imaging using cytoarchitectonic maps. NeuroImage. 2006 Aug 15;32(2):570–82.

Eickhoff SB, Paus T, Caspers S, Grosbras M-H, Evans AC, Zilles K, et al. Assignment of functional activations to probabilistic cytoarchitectonic areas revisited. NeuroImage. 2007 Jul 1;36(3):511–21.

Eickhoff SB, Stephan KE, Mohlberg H, Grefkes C, Fink GR, Amunts K, et al. A new SPM toolbox for combining probabilistic cytoarchitectonic maps and functional imaging data. NeuroImage. 2005 May 1;25(4):1325–35.

Green MJ, Cahill CM, Malhi GS. The cognitive and neurophysiological basis of emotion dysregulation in bipolar disorder. J Affect Disord. 2007 Nov;103(1–3):29–42.

Gu Z, Gu L, Eils R, Schlesner M, Brors B. circlize Implements and enhances circular visualization in R. Bioinforma Oxf Engl. 2014 Oct;30(19):2811–2.

Henson RK. The Logic and Interpretation of Structure Coefficients in Multivariate General Linear Model Analyses. 2002;

Krishnan A, Williams LJ, McIntosh AR, Abdi H. Partial Least Squares (PLS) methods for neuroimaging: a tutorial and review. NeuroImage. 2011 May 15;56(2):455–75.

Le Floch E, Guillemot V, Frouin V, Pinel P, Lalanne C, Trinchera L, et al. Significant correlation between a set of genetic polymorphisms and a functional brain network revealed by feature selection and sparse Partial Least Squares. NeuroImage. 2012 Oct 15;63(1):11–24.

McIntosh AR, Lobaugh NJ. Partial least squares analysis of neuroimaging data: applications and advances. NeuroImage. 2004;23 Suppl 1:S250-263.

McIntosh AR, Mišić B. Multivariate statistical analyses for neuroimaging data. Annu Rev Psychol. 2013;64:499–525.

Moberget T, Alnæs D, Kaufmann T, Doan NT, Córdova-Palomera A, Norbom LB, et al. Cerebellar grey matter volume is associated with cognitive function and psychopathology in adolescence [Internet]. Neuroscience; 2018 Mar. Available from: http://biorxiv.org/lookup/doi/10.1101/288134

Monteiro JM, Rao A, Shawe-Taylor J, Mourão-Miranda J, Alzheimer's Disease Initiative. A multiple hold-out framework for Sparse Partial Least Squares. J Neurosci Methods. 2016 15;271:182–94.

Sherry A, Henson RK. Conducting and interpreting canonical correlation analysis in personality research: a user-friendly primer. J Pers Assess. 2005 Feb;84(1):37–48.

Smith SM, Nichols TE, Vidaurre D, Winkler AM, Behrens TEJ, Glasser MF, et al. A positive-negative mode of population covariation links brain connectivity, demographics and behavior. Nat Neurosci. 2015 Nov;18(11):1565–7.

Xia CH, Ma Z, Ciric R, Gu S, Betzel RF, Kaczkurkin AN, et al. Linked dimensions of psychopathology and connectivity in functional brain networks. Nat Commun. 2018 Aug 1;9(1):3003.

Zohar J, Nutt DJ, Kupfer DJ, Moller H-J, Yamawaki S, Spedding M, et al. A proposal for an updated neuropsychopharmacological nomenclature. Eur Neuropsychopharmacol J Eur Coll Neuropsychopharmacol. 2014 Jul;24(7):1005–14.

Zohar J, Stahl S, Moller H-J, Blier P, Kupfer D, Yamawaki S, et al. A review of the current nomenclature for psychotropic agents and an introduction to the Neuroscience-based Nomenclature. Eur Neuropsychopharmacol J Eur Coll Neuropsychopharmacol. 2015 Dec;25(12):2318–25.

Zöller D, Schaer M, Scariati E, Padula MC, Eliez S, Van De Ville D. Disentangling resting-state BOLD variability and PCC functional connectivity in 22q11.2 deletion syndrome. NeuroImage. 2017 Apr 1;149:85–97.
